# The revolution will be hard to evaluate: How co-occurring policy changes affect research on the health effects of social policies

**DOI:** 10.1101/2020.10.02.20205971

**Authors:** Ellicott C. Matthay, Erin Hagan, Spruha Joshi, May Lynn Tan, David Vlahov, Nancy Adler, M. Maria Glymour

**Affiliations:** Center for Health and Community, University of California, San Francisco, San Francisco, California; University of Minnesota School of Public Health, Department of Epidemiology and Community Health, Minneapolis, Minnesota; Yale University School of Nursing, New Haven, Connecticut

## Abstract

Extensive empirical health research leverages variation in the timing and location of policy changes as quasi-experiments. Multiple social policies may be adopted simultaneously in the same locations, creating co-occurrence which must be addressed analytically for valid inferences. The pervasiveness and consequences of co-occurring policies have received limited attention. We analyzed a systematic sample of 13 social policy databases covering diverse domains including poverty, paid family leave, and tobacco. We quantified policy co-occurrence in each database as the fraction of variation in each policy measure across different jurisdictions and times that could be explained by co-variation with other policies (R^2^). We used simulations to estimate the ratio of the variance of effect estimates under the observed policy co-occurrence to variance if policies were independent. Policy co-occurrence ranged from very high for state-level cannabis policies to low for country-level sexual minority rights policies. For 65% of policies, greater than 90% of the place-time variation was explained by other policies. Policy co-occurrence increased the variance of effect estimates by a median of 57-fold. Co-occurring policies are common and pose a major methodological challenge to rigorously evaluating health effects of individual social policies. When uncontrolled, co-occurring policies confound one another, and when controlled, resulting positivity violations may substantially inflate the variance of estimated effects. Tools to enhance validity and precision for evaluating co-occurring policies are needed.

## INTRODUCTION

Evaluating the health effects of social policies is critical to researchers, funders, and decision-makers seeking to promote healthful, evidence-based programs. Study designs such as differences-in-differences and panel fixed effects (1), which exploit variation in the timing and location of policy changes, have the potential to deliver causal inferences. Changes in health outcomes that are tied to the jurisdictions and times at which a particular policy is adopted can be used to isolate the causal effect of the policy (1). Empirical health research on social policies using these methods has grown rapidly and yielded influential findings in recent years in epidemiology and other fields (2–4).

One major concern with study designs that leverage variation in the timing and location of policy changes is that co-occurrence of policies can render it difficult to separately identify the causal effects of each policy. Isolating individual policy effects is crucial for delivering evidence to decision-makers on whether or not to adopt a policy. Yet multiple related policies are often adopted or implemented in the same jurisdiction simultaneously or in quick succession, rendering it difficult to isolate the effect of one policy from the other. For example, a government that moves to overhaul its social safety net is likely to change multiple welfare-related policies in a single wave of legislative changes (5). Consequently, bundles of related policies, selected to address a particular set of health or social priorities and thus with similar potential health effects, are adopted concurrently, creating co-occurring policies.

Co-occurring policies confound one another. Thus, if the co-occurring policies are relevant to the health outcome of interest, failing to account for co-occurring policies can severely bias estimated effects of specific social policies. For example, if an effective policy A and an ineffective policy B are routinely adopted as a set, and their true effects are unknown, when researchers analyze effects of policy B without accounting for policy A, findings are likely to spuriously indicate that policy B is effective. Yet if jurisdictions typically adopt both policies together, adjustment for policy A to isolate the effect of policy B can lead to imprecise or unstable estimates and bias resulting from data sparsity (6–8). In extreme cases, estimates may be severely biased, undefined, or rely entirely on extrapolation because there is no independent variation in the policy of interest (6).

Strong confounding and consequent data sparsity arising from co-occurring policies can be conceptualized as lack of common support in the data, also known as a violation of the “positivity assumption” (9). Lack of positivity implies that some confounder strata do not have variation in the exposure—for example, because places and times with the confounding policy always adopt the policy of primary interest (the “index” policy). A rich literature exists on the problem of positivity and the use of propensity scores to assess and address it (e.g. by restricting to units that are “on-support”) (9–14). However, several aspects of the policy co-occurrence problem make it important to consider separately from positivity issues that arise with other exposures. First, due to the nature of policymaking (5), the levels of co-occurrence among policy variables may be far greater than those typically observed in non-policy studies (15, 16). For example, governments adopt similar policies at similar times in part because they are responding to the desires and values of their constituents. Second, the most relevant analytic solutions may be distinct. For example, analytic solutions such as data-adaptive parameters (17, 18) that rely on large sample sizes may not be feasible for policy studies that are typically based on a small, fixed set of jurisdictions. Meanwhile, stronger theories or substantive knowledge about the mechanisms by which a particular social policy operates could guide analyses leveraging mediating variables for causal effect estimation (19). For example, how education policy affects educational attainment may be better understood than how educational attainment affects health. Furthermore, if a set of policies are always adopted together, then modifying the exposure definition to encompass both policies and evaluating their combined effect may be the most policy-relevant research question, as opposed to attempting to disentangle their individual effects. Re-conceptualizing the exposure in this way may be less relevant to research in other substantive areas. Thus, the policy co-occurrence problem presents unique challenges and potential analytic solutions beyond typical confounding.

Characterizing the extent and impact of policy co-occurrence is a crucial step for the development of rigorous evidence on social policy effects. Yet, to our knowledge, no epidemiologic research has directly addressed this issue. Prior applied studies of social policies in fields including epidemiology, economics, and political science have acknowledged the issue by critiquing existing policy studies or, in some cases, applying solutions—e.g. studying aggregate measures of policy stringency (20–22). Similar methodological challenges have arisen in environmental epidemiology when studying correlated and multipollutant exposures, but the emphasis of this research has been on identifying analytic solutions appropriate for pollutant measures, rather than on quantifying the extent of the problem (7,8,23). To our knowledge, no prior studies have examined how frequently related policies co-occur, a necessary step to lay the foundation for rigorous analytic solutions. For researchers aiming to estimate individual policy effects, guidance is needed on how to evaluate whether the impacts of policy co-occurrence on estimation are likely to undermine the study. In some cases, the challenge of co-occurring policies may require a modified analytic approach or even altering the research question.

In this paper, we addressed these gaps by proposing and applying an approach to assess the extent of policy co-occurrence and to quantify the impact of policy co-occurrence on the precision of effect estimates for individual policies. Using 13 exemplar social policy databases covering diverse domains, we visually depicted and quantified the extent of policy co-occurrence in each database, and use simulations to estimate impacts on precision. This paper illustrates a novel method that can be used in applied research to determine when policy co-occurrence is so severe that alternative analytic approaches are needed.

## METHODS

### Overview

We developed a systematic sample of social policy databases covering diverse health-related domains that capture measures of policy adoption or implementation across jurisdictions and time. To evaluate the extent and impacts of policy clustering, we applied three analyses to each database. First, we visualized the degree of policy co-occurrence in each database by plotting heatmaps of pairwise correlations among the measured policies. Second, building on the positivity literature, we quantity the overall degree of co-occurrence in each database as the amount of variability in each policy measure across jurisdictions and time that could be explained by the other policy measures in the same database. This indicates how much independent variation remained with which to study the policy of interest. Finally, we used simulations to estimate the impacts of policy co-occurrence on precision by comparing the variance of estimated effects given the observed co-occurrence compared to the variance if all policies were adopted independently.

### Database identification

We sought to characterize the extent of policy co-occurrence across diverse social policy domains. Because no registry of all available social policy databases exists, we identified an exemplar set by evaluating contemporary research on social policies and health, and selecting domain-specific policy databases corresponding to those studies.

We identified all studies of social policies published in 2019 in top medical, public health, and social science journals, emphasizing general-topic journals that publish research on the health effects of social policies: *Journal of the American Medical Association*, *American Journal of Public Health, American Journal of Epidemiology, New England Journal of Medicine*, *Lancet*, *American Journal of Preventive Medicine, Social Science and Medicine, Health Affairs, Demography,* and *American Economic Review.* After these journals had been selected, we asked a convenience sample of 66 researchers from diverse disciplines to rank relevant journals. Responses confirmed that our selected journals reflect common perceptions of most relevant venues for research on the health effects of social policies (detailed results in Appendix: “Survey assessing relevant journals for inclusion”; Appendix Table 1).

We identified original, empirical studies aiming to estimate the causal effects of one or more social policies on health-related outcomes in any country, state, or locality (areas, neighborhoods, or sub-state units such as counties or cities). Although the definition of “social policies” varies across the literature, *a priori* we defined “social policy” to mean any non-medical, population-based or targeted policies that are adopted at a community or higher level, and hypothesized to affect health or health inequalities via changes in social or behavioral determinants. *A priori*, we defined health-related outcomes broadly, to include morbidity, mortality, health conditions, and factors such as smoking, homelessness, and sales of unhealthy products. Given our focus on social interventions, we excluded studies that pertained to health care, health insurance, interventions delivered in the clinical setting, medications, or medical devices, including studies of the Affordable Care Act or Medicaid expansion. For reproducibility, additional detail is presented in the Appendix (“Social policy study inclusion and exclusion criteria”). An independent analyst reviewed a subset of candidate articles to confirm that our strategy to identify relevant papers was reproducible. Concordance between reviewers upon initial review was 90% (for details, see Appendix “Assessment of inter-rater reliability for inclusion and exclusion).

For each social policy study, we identified any corresponding quantitative databases capturing the content, locations, and times of adoption of the index policy and related policies in the same domain. We searched the scientific literature; websites of domain-relevant research institutions, scientific centers, and organizations; and the internet to identify relevant, publicly available databases. We also asked the authors of each index social policy study for policy database recommendations. When possible, we included databases provided on request from individual investigators. If more than one policy database was available, we selected the one that was most amenable to this analysis: first, the database requiring the least data cleaning or manipulation (i.e., panel data structure and variables coded); then, among those remaining, the database with the greatest clarity of variable definitions, followed by the least missingness, and most comprehensiveness (number of policies and time points). We excluded domains for which we could not identify or access any corresponding database. Figure 1 presents information on the number of articles considered, studies and corresponding databases included in the final sample, and studies and databases excluded. Additional detail is presented in the Appendix (“Database selection”).

**Figure 1:**
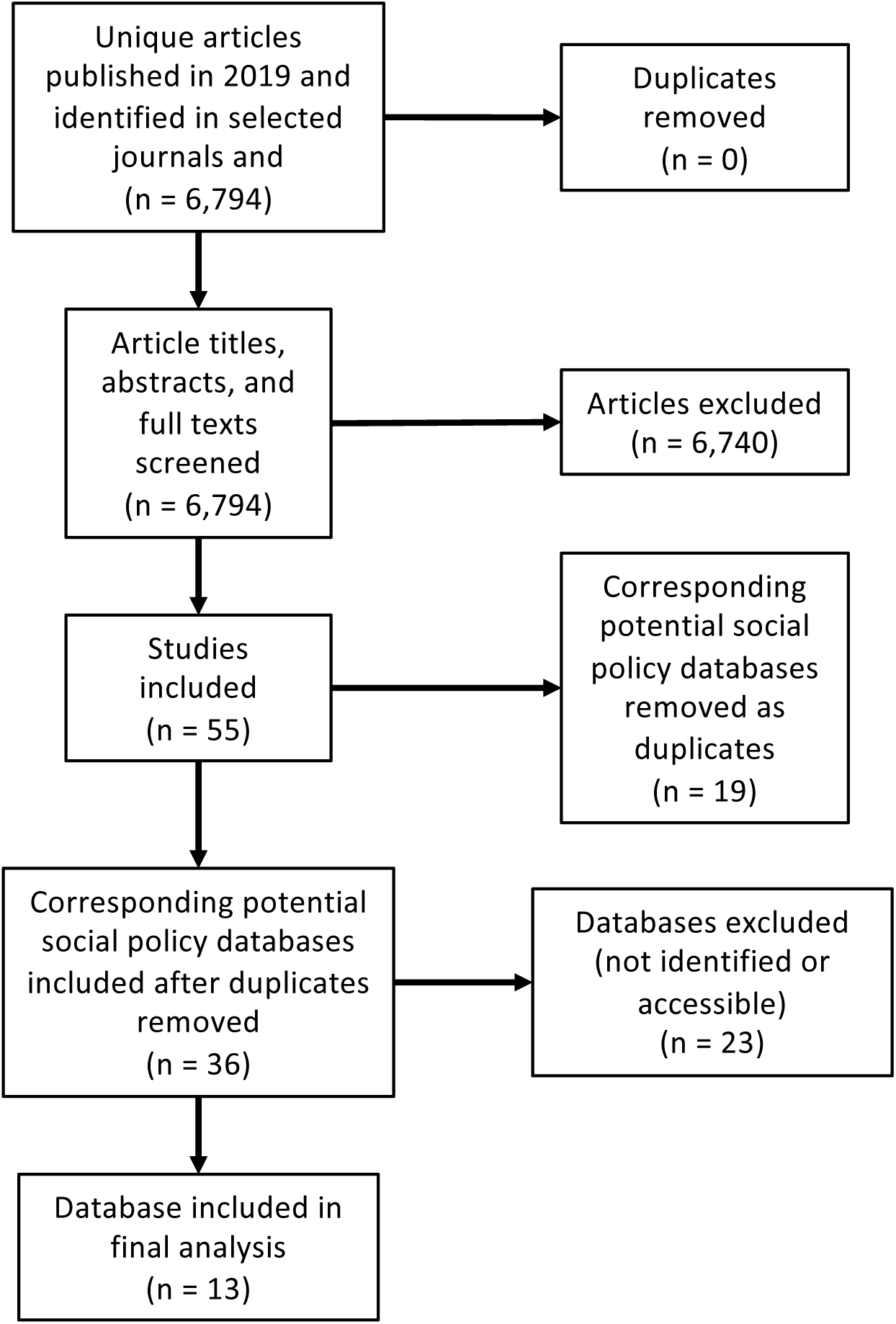
Flowchart of social policy studies and databases included and excluded. Legend: “Corresponding social policy databases that are duplicated” refers to the situation where multiple social policy studies meeting inclusion criteria corresponded to the same database. For example, there were several studies of the impacts of the Supplemental Nutrition Assistance Program (SNAP).

### Database coding

Each database is formatted with one row per jurisdiction (country, state, or locality) and time period (month or year), and one column per policy measure. The types of policy information varied across databases. Some included exclusively binary indicators of policy adoption while others provided information on benefit generosity, implementation, access, and/or scope (e.g. number of Supplemental Nutrition Assistance Program (SNAP) participants by state and year). We included all available policy measures for the heatmaps (see below). For subsequent analyses, when multiple measures of the same policy were available (e.g. year of adoption and number of participants), we selected the measure used in the publication in the original search which invoked the policy, if relevant, or the measure we judged to be the most representative. Some policies were subordinate to umbrella policies. For example, provisions regulating cannabis delivery services are only applicable in jurisdictions where recreational cannabis is legal. For jurisdictions and times in which the umbrella policy was not active, we included these observations in the analysis and coded provisions conditional on that umbrella policy to 0. Additional details are provided in the Appendix (“Database coding”).

### Statistical analysis

First, to visually depict policy co-occurrence in each database, we plotted heatmaps of the Pearson’s correlation matrix for each pairwise combination of policy measures (hereafter, “heatmaps”). Although numerous measures are appropriate, we selected Pearson’s correlation because it is common, intuitive, and accommodates continuous-continuous, continuous-binary, and binary-binary variable comparisons. Although the distribution of Pearson’s correlation between continuous and binary variables is constrained, this constraint is appropriate in this context.

Second, we assessed the degree of unique variation available to estimate individual policy effects, when considering each individual policy while controlling for all others. To do this, we estimated an R^2^ in regression models of each policy regressed on the set of all other policies in the same database. We modeled continuous policy variables using linear regression and used R^2^ adjusted for the number of predictor variables. We modeled binary policy variables using logistic regression and used McFadden’s pseudo R^2^ (24). For both types of regression, we included all predictor policy variables in the database as main terms. This step quantified the amount of variability in each policy across jurisdiction-times that could be explained by the other policy measures and results in a distribution of R^2^ values—one for each policy in each database. This step is also conceptually very similar to estimating propensity scores to assess positivity, except that it accommodates continuous exposure variables.

Third, we estimated the impacts of policy co-occurrence on precision using simulations. For each policy measure, in each policy database, we applied the following procedure:

Step A: Assign a simulated outcome of N observations, where N is the number of jurisdiction-periods in the policy database (Table 1). To simulate the outcome, we assumed (a) a random normal distribution with mean 100 and standard deviation 5; (b) a null effect of the index policy on the outcome (because using an alternative would not substantively affect the results); and (c) 10% of the variance of the outcome was explained by a randomly selected non-index policy (the “explanatory policy”). We incorporate this last component because the precision of the estimated effect of the index policy depends on the proportion of the variance in the outcome that is explained by the other variables in the model. Because large-scale social programs are recognized to have small individual-level effects (25, 26), we considered 10% explained to be optimistic in the setting of the health effects of social policies. We assumed no other confounding was present.

**Table 1:**
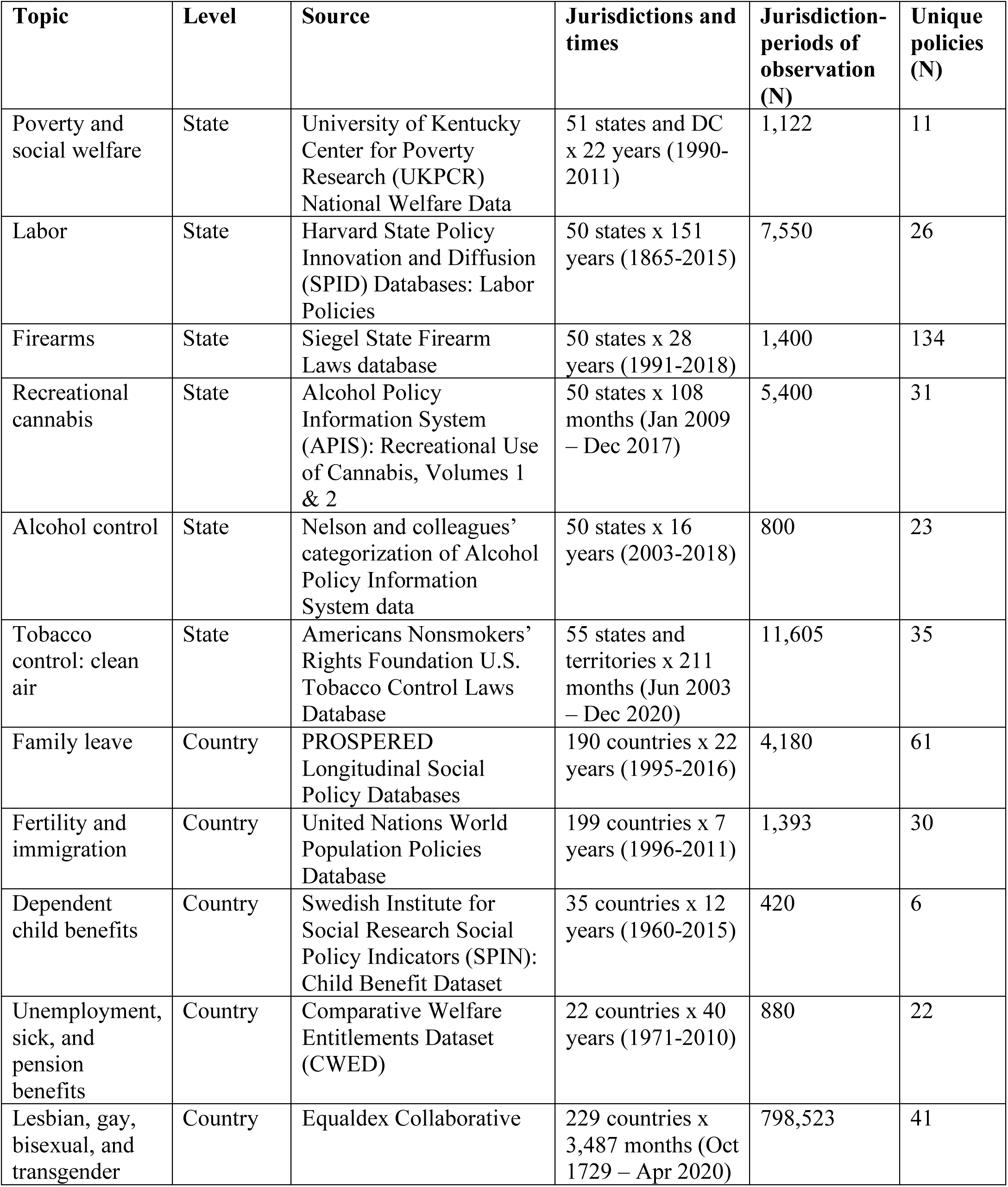
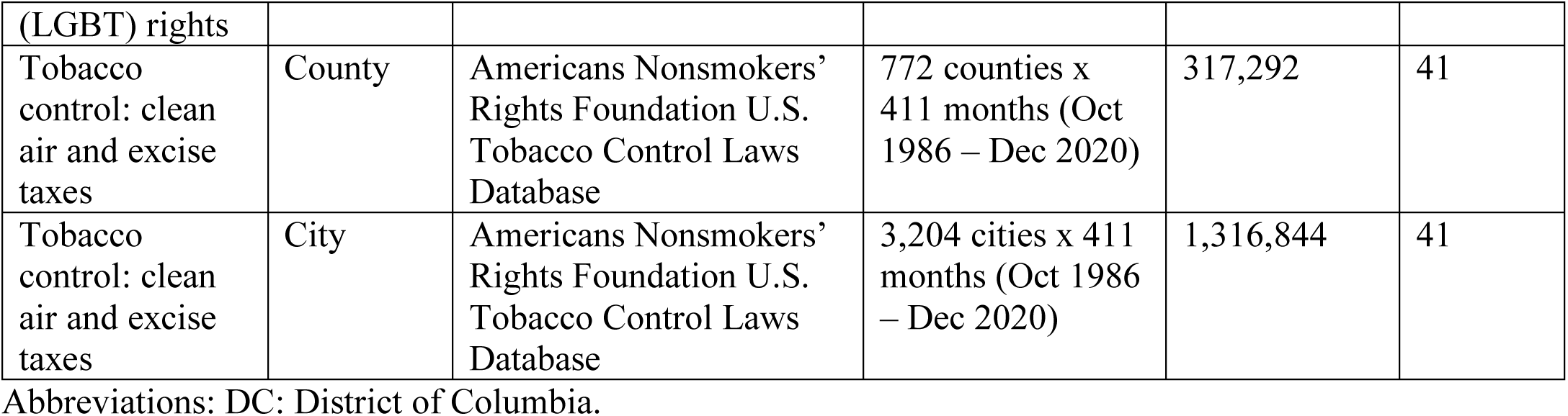
Characteristics of social policy databases included in systematic sample

Step B: Apply a linear regression, modeling the simulated outcome as a function of the index policy, the non-index policies, jurisdiction fixed effects, and time fixed effects. From this regression, record the variance of the regression coefficient corresponding to the effect estimate of the index policy (variance = (standard error)^2^). This was the variance in the real-world, co-occurring, data.

Step C: To estimate the variance if there were no co-occurrence, randomly redistribute the values of the all policy measures across jurisdictions and time (i.e. for each policy measure, randomly shuffling among the rows of the database). This process preserves the overall mean and variance of each policy measure but eliminates systematic co-occurrence.

Step D: Apply the same regression model as in Step B to the redistributed policy data and record the variance of the effect estimate of the index policy.

Step E: Take the ratio of the variance of the effect estimate of the index policy, under the real-world policy regime (derived in Step B) versus under the randomly redistributed regime (derived in Step D) (ratio = variance_StepB_/variance_StepD_). This ratio is an estimate of the variance inflation due to policy co-occurrence.

We conducted Steps A-E 1,000 times for each policy measure in each database, resulting in a set of estimates of the variance inflation. We summarized the variance inflation due to policy co-occurrence for each database by stacking all the variance inflation estimates for all the policy measures in that database and plotting their distribution. We summarized the variance inflation due to policy co-occurrence overall by stacking all the variance inflation estimates for all policy measures in all databases and calculating their summary statistics.

All analyses were conducted using R version 3.6.2.

## RESULTS

We identified 55 studies evaluating links between social policies and health that met our inclusion criteria (27–82), amongst which there were 36 unique policies or databases invoked, and 13 social policy databases that could be identified and accessed (Appendix Table 1; Table 1). Studies included, for example, a panel data analysis of the impacts of changes in the level and duration of paid maternity leave on fertility, workforce participation, and infant mortality across 18 African and Asian countries (37) and a synthetic control evaluation of the effect of raising state-level beer excise taxes on young adult firearm homicides (65).

The sample of 13 identified social policy databases (83–95) (Table 1) included 5 country-level databases, 6 state-level databases, and 2 local-level databases. Domains included poverty and social welfare; family and child welfare; worker welfare; pensions; unemployment; fertility; immigration; LGBT (lesbian, gay, bisexual, and transgender) rights; firearms; alcohol; tobacco; and recreational cannabis. The number of unique policies per database ranged from 6 to 134. Some databases had multiple umbrella policies while others focused exclusively provisions relating to one umbrella policy. For example, the PROSPERED database included overarching policies and specific provisions for breastfeeding breaks, child health leave, family leave, maternity leave, parental leave, paternity leave, and sick leave; while the recreational cannabis policy database focused exclusively on provisions for states where recreational cannabis is legal (e.g. retail sales taxes).

### Visualizing policy co-occurrence

The degree of policy co-occurrence varied by database (Figures 2 and 3; Appendix Figures 1-11). Across the 13 databases, Figure 2 shows an example of intermediate degrees of co-occurrence: amongst unemployment, sick leave, and pension benefits policies across 40 years in 22 countries. Figure 3 displays an example of high levels of co-occurrence: amongst recreational cannabis policies across 108 months in 50 states. Darker colors indicate higher degrees of co-occurrence. Because the correlations are calculated on panel data at the level of the jurisdiction and time unit, higher correlations indicate that jurisdictions which adopt one policy are more likely to adopt the other (or vice versa) and that the policies are likely to be adopted in closer temporal succession.

**Figure 2:**
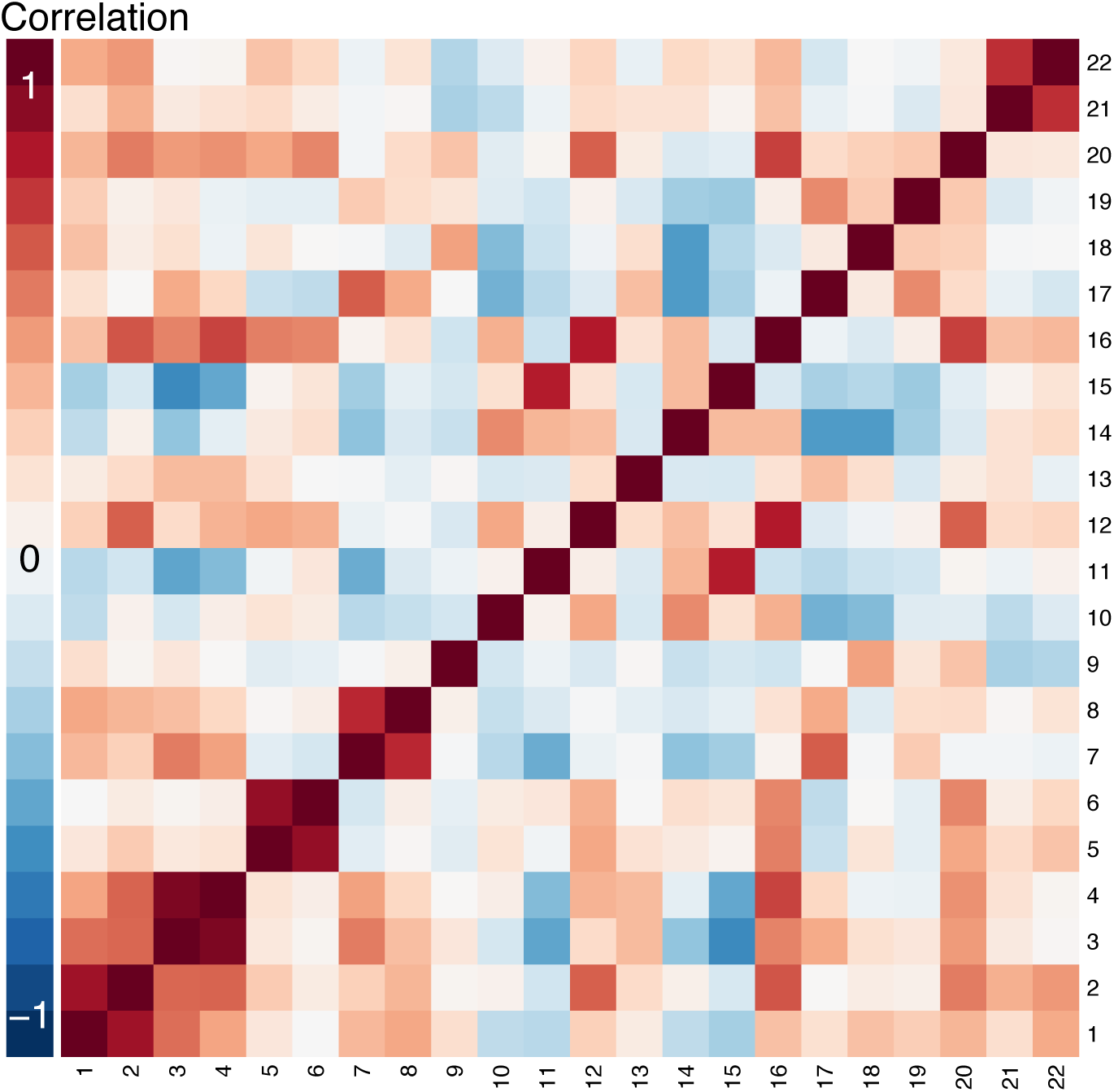
Heatmap of correlations among unemployment, sick, and pension benefits policy measures across country-years, 22 countries, 1971-2010. Legend: High degrees of positive and negative correlation are indicated by the darkest red and blue colors, respectively. Policies: 1: Unemployment insurance replacement rate for single individual living alone. 2: Unemployment insurance replacement rate for single-income family of four. 3: Sickness insurance replacement rate for single individual living alone. 4: Sickness insurance replacement rate for single-income family of four. 5: Minimum pension replacement rate for single individual living alone. 6: Minimum pension replacement rate for single-income family of four. 7: Standard pension replacement rate for single individual living alone. 8: Standard pension replacement rate for single-income family of four. 9: Unemployment insurance qualification period (weeks). 10: Unemployment insurance duration of benefits (weeks). 11: Unemployment insurance waiting period (days). 12: Unemployment insurance coverage (% of labor force insured). 13: Sickness insurance qualification period (weeks). 14: Sickness insurance duration of benefits (weeks). 15: Sickness insurance waiting period (days). 16: Sickness insurance coverage (% of labor force insured). 17: Standard years of pension insurance to be considered fully covered. 18: Ratio of employee pension contributions to employer and employee pension contributions. 19: Years of earnings used in pensionable wage calculation. 20: Pension coverage (% of eligible who are receiving). 21: Male retirement age. 22: Female retirement age.

**Figure 3:**
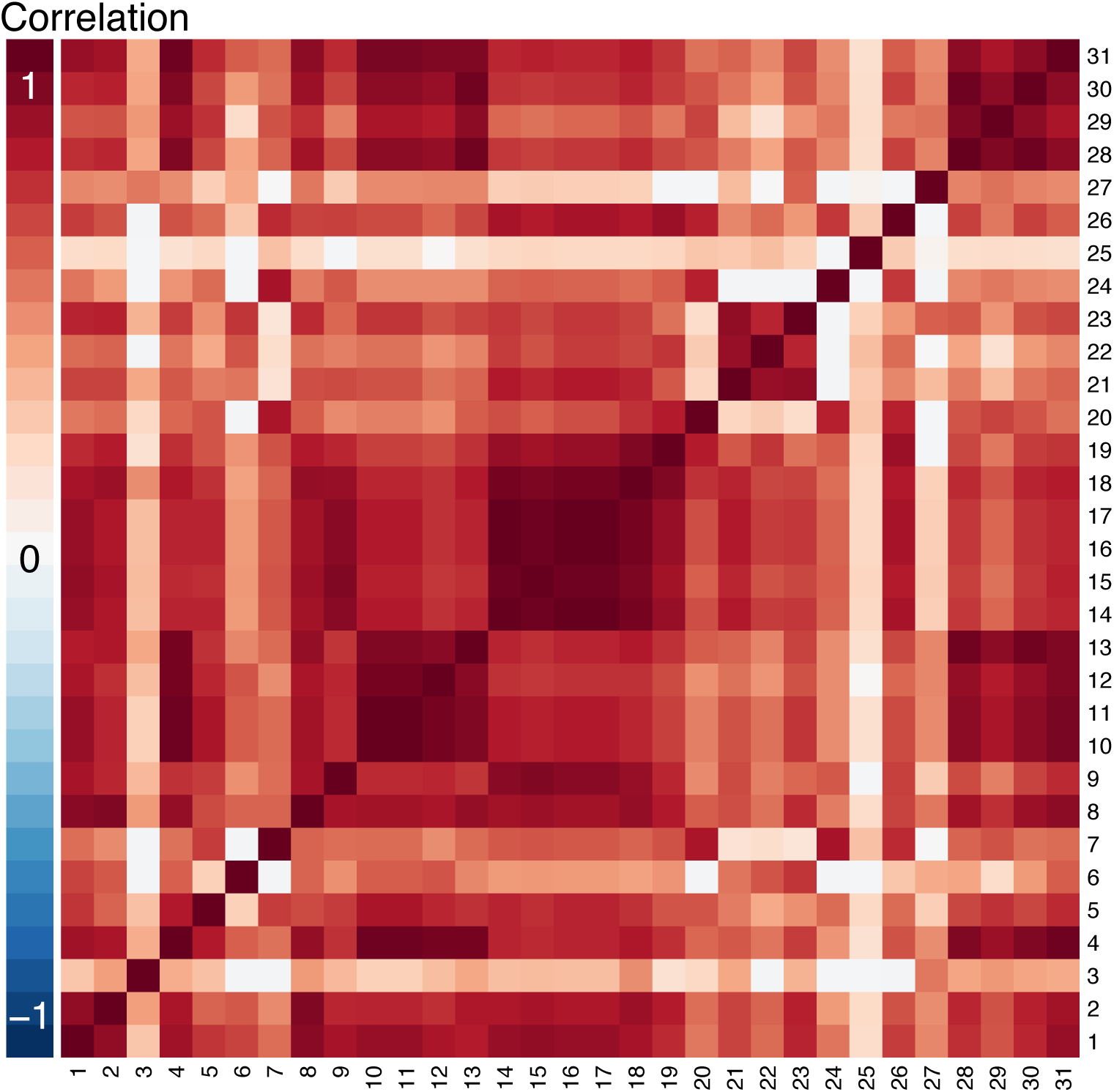
Heatmap of correlations among recreational cannabis policy measures across state-months, 50 states, January 2009 – December 2017. Legend: High degrees of positive and negative correlation are indicated by the darkest red and blue colors, respectively. Policies: 1: Any price controls. 2: Any cultivation restrictions. 3: Allows outlets for on-premise consumption. 4: Allows outlets for off-premise consumption. 5: Any taxes on producers. 6: Retail taxes greater than 15%. 7: Vertical integration prohibited. 8: Tracking system required. 9: Delivery prohibited. 10: Products permitted: edibles. 11: Products permitted: infused products. 12: Products permitted: tinctures. 13: Products permitted: concentrates. 14: Warning labels required: pregnancy. 15: Warning labels required: breastfeeding. 16: Warning labels required: child access. 17: Warning labels required: impairment of driving. 18: Warning labels required: amount of THC. 19: Warning labels required: presence of cannabis or THC. 20: Warning labels required: serving size. 21: Warning labels required: other. 22: Warning labels required: 2 hours to feel effects. 23: Packaging requirements: child resistant. 24: Packaging requirements: child proof. 25: Packaging requirements: tamper evident. 26: Packaging requirements: dose-limited. 27: Packaging requirements: other. 28: Underage possession prohibited. 29: Underage consumption prohibited. 30: Underage purchase prohibited. 31: Allows both state and local cannabis control.

State cannabis policies displayed the highest co-occurrence (median absolute correlation across all pairwise policy combinations: 0.65; 4 policies perfectly aligned) while national LGBT rights policies showed the lowest co-occurrence (Appendix Figure 8; median correlation: 0.04; no policies perfectly aligned). For example, states that restrict what recreational cannabis products can be sold for retail sale also tend to tax retail cannabis sales, while countries that allow same-sex marriage were relatively independent of countries that ban LGBT-related employment discrimination. Tobacco policies at the state, county, and city jurisdiction levels showed similar degrees and patterns of co-occurrence among policies—for example, comprehensive clean air laws for bars and comprehensive clean air laws for restaurants were frequently co-occurred at the state, county, and city levels; Appendix Figures 9-11).

Most policy measures were positively correlated, but we also found pockets of negative correlations. For example, country-years with child tax credits tended not to have child tax allowances (Appendix Figure 1). The heatmaps also reveal groups of co-occurring and independent policies. For example, labor policies requiring licensing for different professions frequently co-occurred, but this set was relatively independent of policies regarding collective bargaining and minimum wages (Appendix Figure 5).

### Quantifying policy co-occurrence

Most of the variability in policy measures across jurisdiction-times was explained by the other policies in the same database. Figure 4 displays the distributions of R^2^ values: the higher the R^2^, the less unique variation there is for an individual policy, to a maximum of 1.0. The impacts of policy co-occurrence on identifiability were generally substantial: of all 502 policies considered, 65% had R^2^ values greater than 0.90 when regressed on other policies in the same database. Child benefits had the lowest R^2^ distribution, with a median of 0.19; poverty and social welfare, family leave, fertility/immigration, firearms, cannabis, alcohol, state tobacco control, and county tobacco control policies had R^2^ distributions with medians around 0.9 or higher. In some cases, correlations between predictor policy variables were so strong that the statistical software forced certain variables from the model (frequency reported in Appendix Table 2).

**Figure 4:**
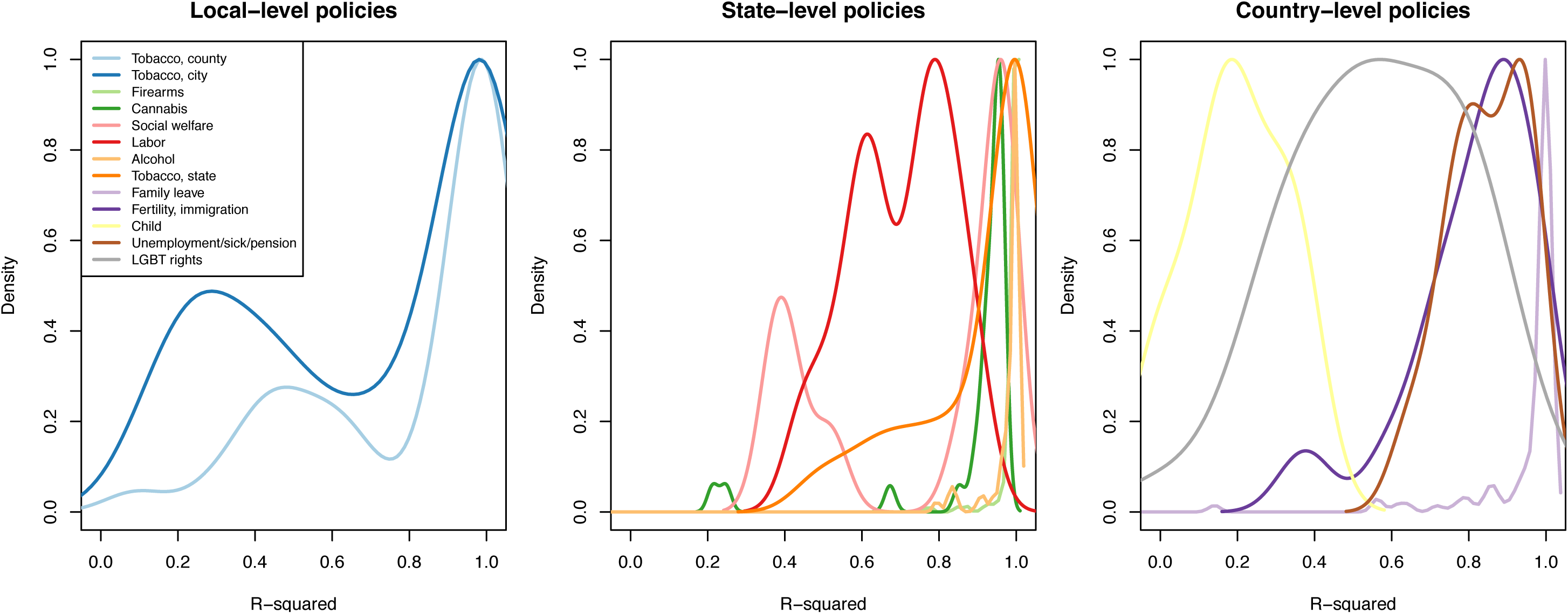
**Distributions of proportions of variability in each policy that is explained by other policies (R^2^) across social policy databases**

### Impacts of policy co-occurrence on precision

Policy co-occurrence substantially reduced the precision of possible effect estimates in all cases (Figure 5). Across policy measures, databases, and simulation iterations, policy co-occurrence effectively increased the variance of effect estimates by a median of 57-fold. Across policies, the lowest degree of variance inflation observed was 7% (median across simulations) for country child tax rebates. For other policies, particularly family leave, variance inflation was so substantial as to render estimates effectively indeterminate. Again, some predictors were dropped from models due to strong correlations with other predictors (Appendix Table 2).

**Figure 5:**
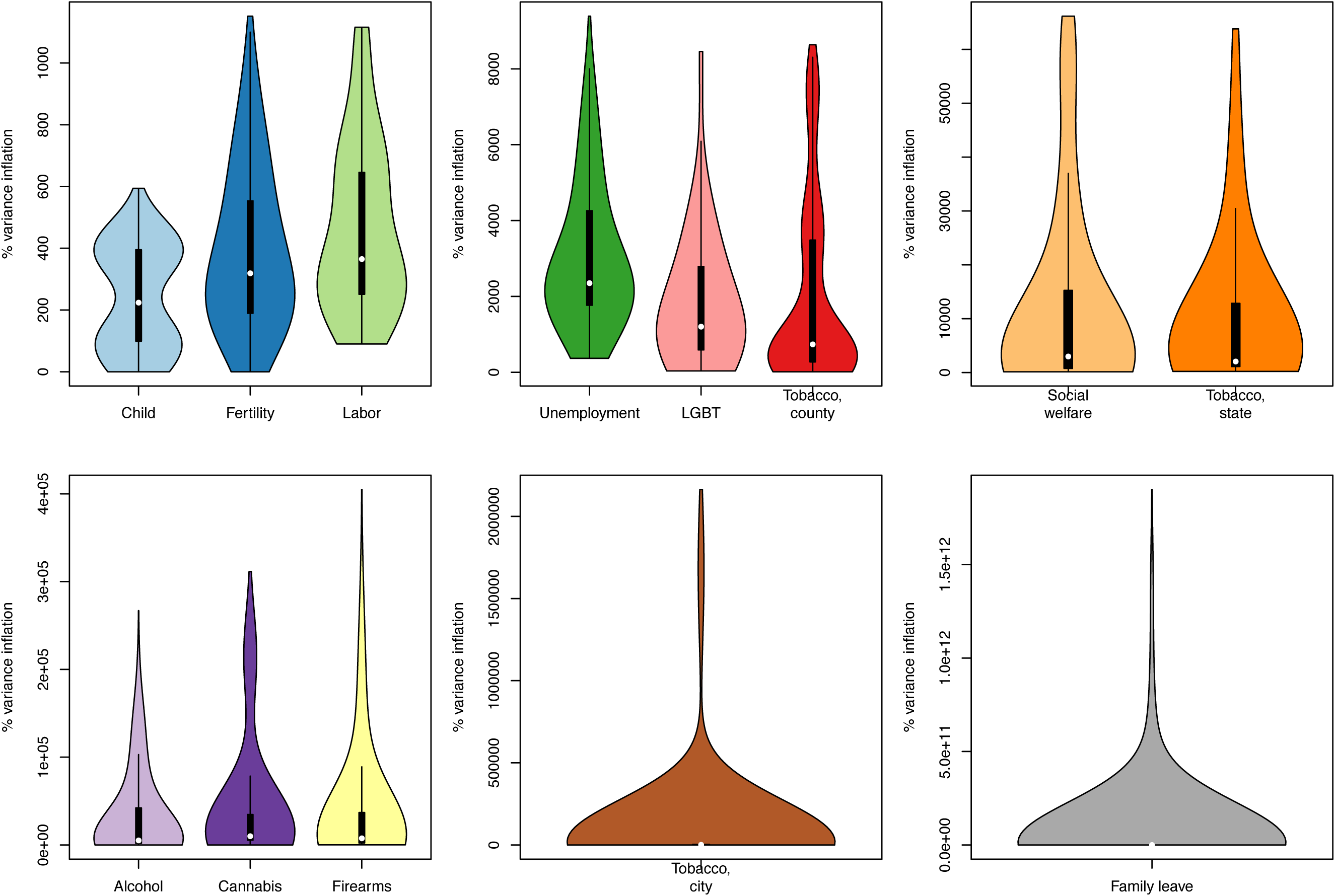
Distributions of simulated impacts of policy clustering on the precision of estimated effects. Abbreviations: LGBT: Lesbian, gay, bisexual, and transgender rights. Fertility: Fertility and immigration policies. Unemployment: Unemployment/sick/pension benefits.

## DISCUSSION

We analyzed 13 social policy databases drawn from contemporary research in top epidemiology, clinical, and social science journals. These exemplar databases represented diverse policy domains, geographies, and time periods to describe the pervasiveness and impacts of policy co-occurrence on estimation of health effects. We found that high degrees of co-occurrence were the norm rather than the exception. For a majority of policies, greater than 90% of the variation across jurisdiction-times was explained by other related policies in the same database. Unbiased studies attempting to isolate individual policy effects must control for these related policies, so for many applications, there may be little independent variation left with which to study the policy of interest. Consistent with this, we found that adequate control for co-occurring policies is also likely to substantially reduce the precision of estimated effects, often so dramatically that informative effect estimates are unlikely to be derived.

### Interpretations

Several factors make the pervasiveness and consequences of policy co-occurrence likely to be even greater than we have estimated. First, we only examined policy co-occurrence within domain-specific databases. Yet social policy changes may happen in multiple domains simultaneously. For health outcomes affected by diverse types of policies (e.g. both unemployment policies and firearm policies may affect suicide rates), researchers must consider policy co-occurrence *across* domains which likely indicate even more severe co-occurrence. Second, each policy database we considered included only one jurisdictional level, but true policy environments involve complex overlays of national, state/province, county, municipal, employer, and/or school policies. Third, we did not incorporate lagged effects or nonlinear relationships between variables. Fourth, policy variables that perfectly or near-perfectly predict one another were dropped from the regression models. Finally, we did not consider the multitude of social, economic, political, or societal factors (e.g. a recession, migration, gentrification), that may also co-occur with policies of primary interest, including changes in social norms, implementation, or enforcement that can be conflated with policy changes. Some such confounders can be controlled with jurisdiction or time fixed effects; measured confounders that are jurisdiction-specific and time-varying could be evaluated using the same methods illustrated here. This is a formidable task; data sharing efforts would facilitate its assessment and handling.

We found that the overall degree of policy co-occurrence varied across databases, ranging from very high for state-level recreational cannabis policies to low for country-level sexual minority rights policies. Several different factors may drive this variation. Our finding that tobacco policies at the state, county, and city levels showed similar degrees and patterns of co-occurrence among similar sets of policies suggests that co-occurrence may be a characteristic of the domain. Political polarization may result in greater co-occurrence for certain policy domains (e.g. firearms) versus others (e.g. family leave). Databases with rarer policies, fewer umbrella policies (e.g. recreational cannabis), or more nested policies (e.g. firearm policies that apply to all guns versus handguns) also tended to have more co-occurrence. Databases with more unique policies also generally had more co-occurrence; with a fixed number of jurisdiction-times of observation, considering more policies creates more opportunities for alignment. Importantly, these patterns highlight that the measured degree of co-occurrence depends not only on the policies themselves but also on the investigator’s choices of policy measures. Further, policies that could be considered alternatives rather than complements (e.g. child tax credits and child tax allowances) co-occurred less frequently and may offer the opportunity for more robust studies of causal impacts. Differences in the ways that policies are adopted across different political systems and different jurisdictional levels may also matter. In our examples, country-level policies appeared to co-occur less frequently than state-level policies, implying that estimating causal effects of country-level policies may be more feasible. Similar considerations apply to the temporal scale of analyses as well: the feasibility of estimating health effects may depend on whether analyses are conducted at the level of the year, month, or even election cycle. Our analysis could not determine which of these factors drives variation in policy co-occurrence; this would be a fruitful area for future research.

### Limitations

Several other limitations of this study must be noted. Certain policy domains were not covered, either because no social policy studies for that domain were sampled, or because no corresponding policy database was identified or accessed. We did not review all potentially relevant journals. Our results may not generalize to policy domains or journals not examined. Our approach also assumes that all the policies in each domain-specific database are relevant to the health outcome of interest; this is plausible for social interventions that likely affect a broad range of health outcomes, but for some outcomes, only a subset of the policies in a database may need to be controlled to isolate the effect of the index policy. Additionally, our approach is only relevant when a database of the relevant policies exists or can be constructed. Developing policy databases is often an arduous task requiring systematic review of legal language. We did not consider the quality of the underlying databases. Our selections serve to illustrate the policy co-occurrence problem, but for applied researchers, the optimal policy database may differ from the one used here. The problem of correlated exposures arises in many domains, including environmental health, and although social policies are distinct in important regards, methods in other domains may nonetheless prove helpful. Furthermore, our analysis did not examine the distinctions between policy adoption, implementation, promulgation, or changes in norms that precede or follow from policy changes, but these considerations are essential in applied policy research.

Finally, data sparsity arising from co-occurring policies can lead to bias not just imprecision. Our simulations did not incorporate this because this type of bias is less relevant to studies of the health effects of social policies, and highly context-specific. Simulation results on the magnitude of bias from positivity violations are therefore unlikely to be generalizable. Specifically, bias arising from positivity problems depends on the estimation method. For methods that rely on modeling the outcome (e.g. with regression), positivity-related bias arises from model-based extrapolation. For methods that involve modeling the exposure mechanism (e.g. propensity score matching, inverse probability of treatment weighting), bias can result from disproportionate reliance on the experiences of a just a few units, or the absence of certain confounder strata (i.e. certain groups are never exposed and thus cannot be weighted to de-bias the estimate). Since our simulations are based on outcome regressions—the most common approach for differences-in-differences, panel fixed effects, and related designs—bias would only arise from model-based extrapolation. However, for the vast majority of policies identified in this study, measures were binary and thus extrapolation cannot occur. For continuous policy measures (e.g. the amount of a tax), model-based extrapolation is possible, but application-dependent. Thus, simulating the potential degrees of bias resulting from model-based extrapolation requires either tenuous generalizations or substantive assumptions about each policy area. We suspect that extremely non-linear relationships that would lead to large extrapolation bias are rare for policy effects, but this remains an open question.

### Implications

Researchers should be cautious when seeking to make causal inferences about the health effects of single social policies using methodological approaches premised on “arbitrary” or quasi-random variation in policies across jurisdictions and time. Not every policy change offers a valid differences-in-differences or panel fixed effects study design. These methods are most compelling when policy implementation is staggered across jurisdictions and dates independently from other policies and for plausibly like-random or arbitrary reasons. For example, there could be differing timing of elections, legislative sessions, “crises” that provoke specific policy changes, or “lottery”-type rollouts. In these cases, such research can be very persuasive, or at least constrain the set of co-occurring policies. Our finding of pervasive policy co-occurrence across numerous databases suggests that many policies do not fit this criterion.

Inadequate control for co-occurring policies or differences in the set of policies controlled may explain surprising or conflicting results in previous studies. Investigators should base interpretations of social policy research on the plausibility that policy adoption is distributed arbitrarily with respect to other uncontrolled policies or social changes, a phenomenon that in reality may be rare. This evaluation should be based on deep content knowledge of law, politics, and society—a compelling argument for involving policymakers in the design and interpretation of studies.

### Potential solutions

We illustrate an approach for researchers to assess whether the effects of individual policies can be estimated. While other simulation-based methods for assessing positivity exist (9), the approach we propose is tailored to the policy co-occurrence problem and facilitates examining how a full set of policies substantively occur together. For a given application, if the heatmap indicates high correlations, and estimated R^2^ values and variance inflation are high, it may be necessary to alter the research question and corresponding analytic approach.

Researchers have applied numerous analytic approaches to address the challenge of highly co-occurring policies, ranging from machine learning algorithms that identify policy measures most strongly related to an outcome of interest to methods that characterize overall policy environments based on expert panels. Relevant methods have been discussed in diverse prior work (see for example (7,8,10,13,23,96–99)). The second paper in this series on policy co-occurrence provides a systematic assessment of available methods. We briefly review three promising analytic options here, and refer the reader to the second paper in this series for more detail.

One approach is to focus on outcomes that are affected by the index policy of interest but not the co-occurring policies. For example, changes in state Earned Income Tax Credits (EITC) co-occur with changes in other social welfare policies (100). Rehkopf and colleagues took advantage of seasonality in the disbursement of EITC cash benefits (typically delivered in February, March, and April), versus benefits without the same seasonal dispersal pattern, to examine the association of EITC with health using a differences-in-differences design (101). By comparing health outcomes that can change on a monthly basis (e.g. health behaviors) for EITC-eligible versus non-eligible individuals in months of income supplementation versus non-supplementation, the authors measured potential short-term impacts of EITC independent of other social welfare policies.

Another approach is to move beyond binary measures of policy adoption to more detailed characterizations—the amount of funding, generosity, participation rate, or population reach of a program; the size of a tax; or the duration of a policy. These measures may co-occur less frequently with related policies, or provide opportunities to examine dose-response effects among jurisdictions adopting the policy. For example, the adoption of certain unemployment benefit policies co-occurs frequently with other social welfare policies across state-years. Researchers have successfully assessed their health impacts by comparing varying levels of unemployment benefit generosity—measured as the total maximum allowable benefit (in US dollars) per bout of unemployment –across states and years (102, 103). Heatmaps like those presented in this study may help researchers to identify specific policy measures that are more independent from related policies.

A final option is to conceptualize policy clusters, instead of individual policies, as exposures. This is promising if policies are typically adopted as a group, as is the case with the large omnibus bills that are increasingly common at the state and federal levels. This approach is also particularly relevant when studying the provisions of a single umbrella policy. For example, for provisions of recreational cannabis legalization, exposure categories based on the overall approach to legalization in one state versus another may be of greater interest than the effects of individual provisions. Similarly, Erickson and colleagues categorized states into four groups based on stringency of the overall alcohol policy environment and found that these categories were associated with levels of past-month alcohol consumption (104). Several options are available to define clusters, including manual selection, hierarchical cluster analysis, latent class analysis, and principal components analysis (105). Heatmaps like those presented here can help inform the selection of appropriate clusters by offering an intuitive visual reference for the likelihood that sets of policies were adopted together. Evaluating situations when each clustering approach might be preferable is a future research direction.

## Conclusions

Overall, our findings suggest that co-occurring policies are a major methodological challenge to rigorously evaluating the health effects of individual social policies. Rigorous study design and interpretation of studies that seek to isolate individual policy effects requires careful attention to co-occurring policies and their impacts on identifiability and precision. Evaluating the health effects of policies is a powerful strategy to address confounding and an important substantive domain, conceptualizing social policies as a natural avenue for translation of epidemiologic findings to public health. Study designs, statistical methods, and data collection efforts to enhance statistical power for evaluating co-occurring policies or to circumvent the co-occurrence are a high priority for future work.

## Data Availability

This study involves only secondary data on social policies that is publicly available online.

## Appendix

### Supplemental Methods

#### Survey assessing relevant journals for inclusion

We developed a short online survey of population health researchers, disseminated through the authors’ professional networks via email, Twitter, and LinkedIn. The survey stated: “We’d like to identify an illustrative set of journals where important research on the health effects of social policies is published. We want journals that are (a) high impact, (b) reflect diverse disciplines, and (c) publish original quantitative research on how social policies influence population health.” For each of 5 disciplines (epidemiology, public health, clinical medicine, sociology/demography, and economics/health policy), we provided 8 candidate journals, informed by the highest ranking general-topic journals in each field across all countries/regions according to Scimago Journal Rankings. We asked respondents to rank the most important journals to include in each of the 5 fields, with the option to skip fields with which the respondent is not familiar. Response options were ordered randomly for each respondent. This survey was deemed not to be human subjects research by the University of California, San Francisco Human Research Protection Program.

We received 66 anonymous responses, with self-identified primary disciplines of epidemiology (27 respondents), public health (15), medicine (7), sociology/demography (9), economics/health policy (7), and other disciplines (7). The table below summarizes the rankings results.

A clear “top journal” for research on the health effects of social policies emerged in each discipline and our study included all of these top-ranking journals. The 5 other journals we included were ranked 2^nd^ or 3^rd^. There were 29 additional journals suggested by respondents, none of which were suggested by more than one or two respondents, except Health Services Research, which was suggested by 4 of 66 respondents. There were 11 additional disciplines suggested by respondents (e.g. geography, social work) but none was suggested by more than one respondent.

**Appendix Table 1:** Top-ranking journals publishing high-impact research on the health effects of social policies, according to a convenience sample of 66 population health reviewers

| **Discipline** | **Journals included in current study** | **Votes for 1<sup>st</sup> place** | **Included in current study** |
| --- | --- | --- | --- |
| Epidemiology | *American Journal of Epidemiology* | 26 | Yes |
|  | *Journal of Epidemiology and Community Health* | 9 |  |
|  | *Epidemiology* | 7 |  |
|  | *International Journal of Epidemiology* | 4 |  |
|  | *Annals of Epidemiology* | 3 |  |
|  | *European Journal of Epidemiology* | 2 |  |
|  | *Epidemiologic Reviews* | 1 |  |
|  | *Journal of Clinical Epidemiology* | 0 |  |
| Public health | *American Journal of Public Health* | 42 | Yes |
|  | *Lancet Public Health* | 8 |  |
|  | *American Journal of Preventive Medicine* | 3 | Yes |
|  | *Morbidity and Mortality Weekly Report* | 3 |  |
|  | *Preventive Medicine* | 1 |  |
|  | *BMC Public Health* | 1 |  |
|  | *Eurosurveillance* | 0 |  |
|  | *Bulletin of the WHO* | 0 |  |
| Medicine | *New England Journal of Medicine* | 24 | Yes |
|  | *Journal of the American Medical Association* | 11 | Yes |
|  | *Lancet* | 9 | Yes |
|  | *PLoS Medicine* | 4 |  |
|  | *British Medical Journal* | 3 |  |
|  | *Science* | 3 |  |
|  | *Annals of Internal Medicine* | 2 |  |
|  | *Nature Medicine* | 2 |  |
| Sociology/demography | *Social Science & Medicine* | 30 | Yes |
|  | *American Sociological Review* | 10 |  |
|  | *Demography* | 6 | Yes |
|  | *American Journal of Sociology* | 4 |  |
|  | *Journal of Health and Social Behavior* | 4 |  |
|  | *Social Forces* | 1 |  |
|  | *Demographic Research* | 0 |  |
|  | *European Sociological Review* | 0 |  |
| Economics/health policy | *Health Affairs* | 38 | Yes |
|  | *Journal of Health Economics* | 8 |  |
|  | *American Economic Review (AER)* | 7 | Yes |
|  | *Milbank Quarterly* | 4 |  |
|  | *Health Policy* | 1 |  |
|  | *American Journal of Political Science* | 0 |  |
|  | *Quarterly Journal of Economics* | 0 |  |
|  | *Journal of Political Economy* | 0 |  |

#### Social policy study inclusion and exclusion criteria

Included:

- Original, empirical studies aiming to make inferences about the causal effects of one or more social policies on health-related outcomes

○ Although the boundaries of “social policies” are fuzzy, we focused on non-medical, population-based or targeted policies that are adopted at a community or higher level and affect social determinants of health or social inequalities in health.
○ We defined health-related outcomes broadly, to include morbidity, mortality, and health conditions, as well as factors related to health outcomes such as smoking, neighborhood availability of health foods, health expenditure, utilization of health services homelessness, and sales of healthy and unhealthy products
- Other inclusions:

○ Economic, welfare, unemployment, family and child-related policies
○ Food and nutrition-related policies
○ Drug and alcohol policies
○ Road safety policies
○ Immigration policies
○ Firearm policies
○ Policies regulation federal school meals

Excluded:

- Studies that pertained to health care, health insurance, interventions delivered in the clinical setting, medications, medical procedures, or medical devices, including studies of preventive or treatment-oriented health services, the Affordable Care Act, and Medicaid expansion
- Systematic reviews and meta-analyses
- Qualitative and ethnographic studies
- Hypothesis-generating studies
- Simulation studies, unless they estimated the effects of existing policies
- Studies that leverage policy changes as an instrument to study the effects of endogenous variables such as educational attainment or income (these studies do not estimate the effects of social policies)
- Other exclusions:

○ Interventions delivered in the clinical setting, even if they are delivering social services (e.g. housing supports)
○ Surveys about potential responses to hypothetical regulations used to project the impacts of future/potential policies
○ RCTs unless used to evaluate and actual, real-world policy
○ Studies of the effects of lawsuits
○ Studies about cancer screening or communicable testing incentives, guidelines, recommendations and policies
○ Mechanisms and structures of health care payments; health care finance mechanisms and models
○ Studies of industrial/macroeconomic policies with productivity, employment, income, or wages as outcomes
○ Studies on health care spending
○ Studies of the health information exchange
○ Millennium development goals
○ Infectious disease harm reduction
○ Mass drug administration
○ Democracy, regime type, or free and fair elections
○ Community-based interventions around medication access and adherence
○ Vaccination policies
○ Federal funding for research
○ Physician shortages
○ Expansions of family planning services
○ Mental health policies, even if they have components that are not about health insurance or health services – e.g. stigma reduction campaigns (still mainly a health services policy)
○ Funding for prevention programs or primary care
○ Incentives for behavior change in Medicaid
○ Moving to Opportunity
○ Government funding earmarked for different purpose but not associated with a particular policy

#### Assessment of inter-rater reliability for inclusion and exclusion

We sought to assess the replicability and reliability of classifying each journal’s 2019 articles as “studies of the health effects of social policies” or not. We recruited one analyst to independently review a random subsample of the journal articles we initially reviewed for inclusion. The analyst’s training background was an MPH in Environmental Health. We provided her with some background on the project, some general training in what constitutes a social policy, and the definitions and inclusion/exclusion criteria provided in the main text and appendix of our paper. We provided the analyst with 31 full-text manuscripts: 10 meeting inclusion criteria and 21 studies not meeting inclusion criteria. The analyst classified 28 of the 31 studies in concordance with our determination of inclusion/exclusion. Conflicts for the three discordant studies were easily resolved with the determination to exclude after further clarifying that a composite index of country-level participation in conflict and individually-initiated abstinence-based fertility control were not measures of social policies, and that the exclusion of instrumental variables studies only applied to studies in which leverage policy changes as an instrument to study the effects of endogenous variables such as educational attainment or income.

#### Database selection

For each social policy study, we identified any corresponding quantitative databases capturing the content, locations, and times of adoption of that index policy and related policies in the same domain. We searched the scientific literature; websites of domain-relevant research institutions, scientific centers, and organizations; and the internet to identify relevant, publicly available databases. We also asked the authors of each index social policy study for policy database recommendations. When possible, we included databases that were provided on request from individual investigators.

If more than one policy database was available, we selected the one that was most amenable to this analysis: first, the one requiring the least data cleaning or manipulation (i.e., panel data structure and variables coded); then, among those remaining, the one with the greatest clarity of variable definitions, followed by the least missingness, and most comprehensiveness (number of policies and time points). For example, the *American Journal of Preventive Medicine* published a January 2019 study on the association of state firearm legislation with intimate partner homicide using the RAND State Firearm Law Database (97); several state firearm law databases exist, and we selected the one developed by Siegel and colleagues because it had the cleanest data, most precise variable definitions, and covered the greatest number of policies (98, 99). The database selected for our analysis was not always the one used in the index policy.

For studies evaluating national-level policies, we treated the corresponding database as the one with country-level panel data, even if the corresponding study did not include data from other countries—for example, if the authors used an interrupted time series design. If we could not find a database that included the location in the index study (e.g. China), we used the best available database covering other geographic units, if one was available (e.g. OECD countries). We excluded studies and domains for which we could not identify or access a corresponding database.

#### Database coding

Each database is formatted with one row per place and time period. Each policy was a separate column indicating the value of the policy measure for the given place and time. The time periods were defined based on the finest resolution available in the data set (in most cases, years). The types and formats of policy information varied across databases. For example, some included exclusively binary indicators of when each policy was adopted while others provided information on benefit generosity, implementation, access, or scope (e.g. number of Supplemental Nutrition Assistance Program (SNAP) participants by state and year). Some databases code laws very specifically (e.g. whether a country child family leave policy requires a minimum employee tenure) while others were more general (e.g. whether a country has any policy to increase immigration). Some included dates of enactment, dates the laws became effective, and dates that the laws were amended, if relevant; we included these as separate policy measures. Most included laws, resolutions, and regulations, but excluded executive orders and determinations resulting from legal cases. We included all available policy measures for the heatmaps. For subsequent analyses, when multiple measures of the same policy were available (e.g. year of adoption and number of participants), we selected the measure used in the publication in the original search which invoked the policy, if relevant, or the measure we judged to be the most representative.

We converted categorical policy variables to a series of dichotomous variables, with “no policy” serving as the reference category. Some policies were subordinate to umbrella policies. For example, provisions regulating cannabis delivery services are only applicable in jurisdictions where recreational cannabis is legal. For jurisdictions and times in which the umbrella policy was not active, provisions conditional on that umbrella policy were coded to 0. We treated two policies where one is nested in the other as separate policy variables. For example, handguns are a type of gun; we treated policies requiring a waiting period on purchases of all handguns and policies requiring a waiting period on purchases of all guns as separate policy variables. Additional details on database coding are provided in the Appendix (“Database coding”).

Some databases simply provided a list of the dates of adoption of a set of policies, leaving ambiguity about how many time-units prior to policy adoption should be included in the analysis. In applied work, this would likely depend on the health outcome, data availability, and number of time units needed to establish the pre-policy outcome trends. For our analyses, when converting these to place-time period datasets, we chose to include three time-units before adoption of the first policy and after adoption of the last policy (when relevant), as similar duration run-in and run-out periods are common in social policy studies. If exact dates of policies were provided, we converted the data to a jurisdiction-month-level panel database. We treated missing data—for example, missingness arising because a country was not an entity in a given year—as missing completely at random. We reported the degree of missingness in Appendix Table 2 and conducted complete case analysis. Some databases lacked information on all policies for all available years; in this case, we restricted the range of years or set of policies included in the final database to maximize the number of state-year-policy data points included.

### Supplemental results

**Appendix Table 1:**
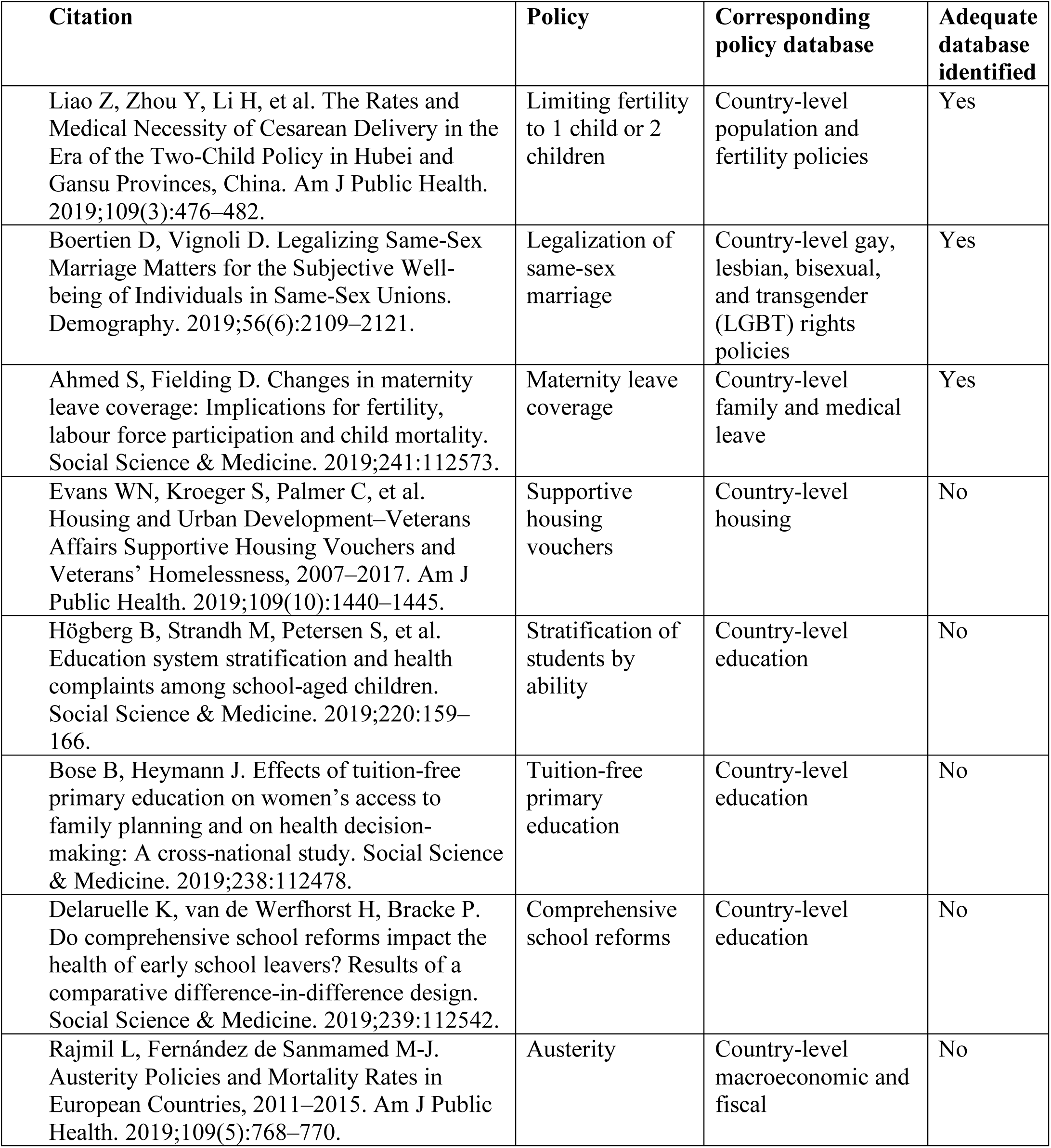
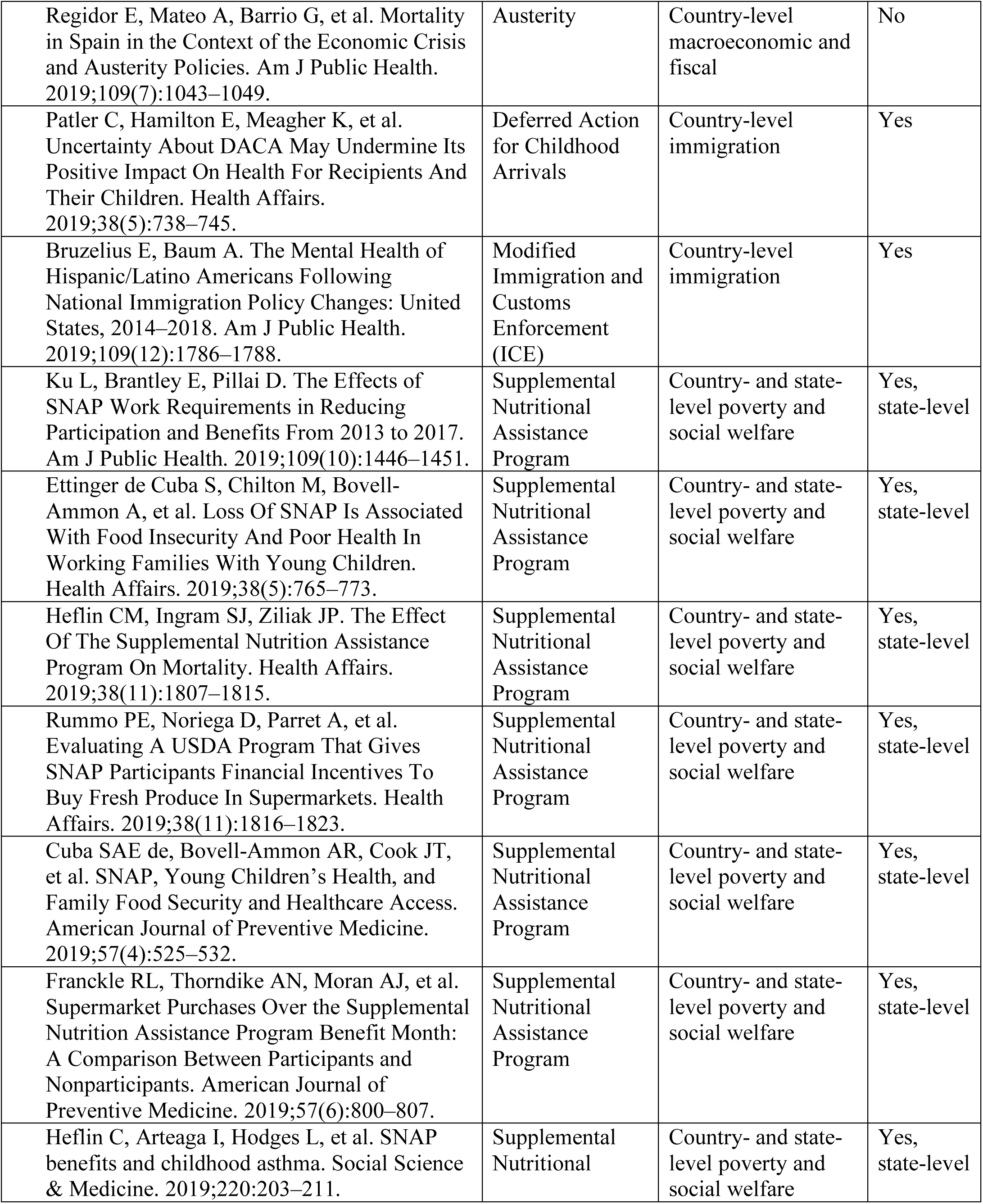
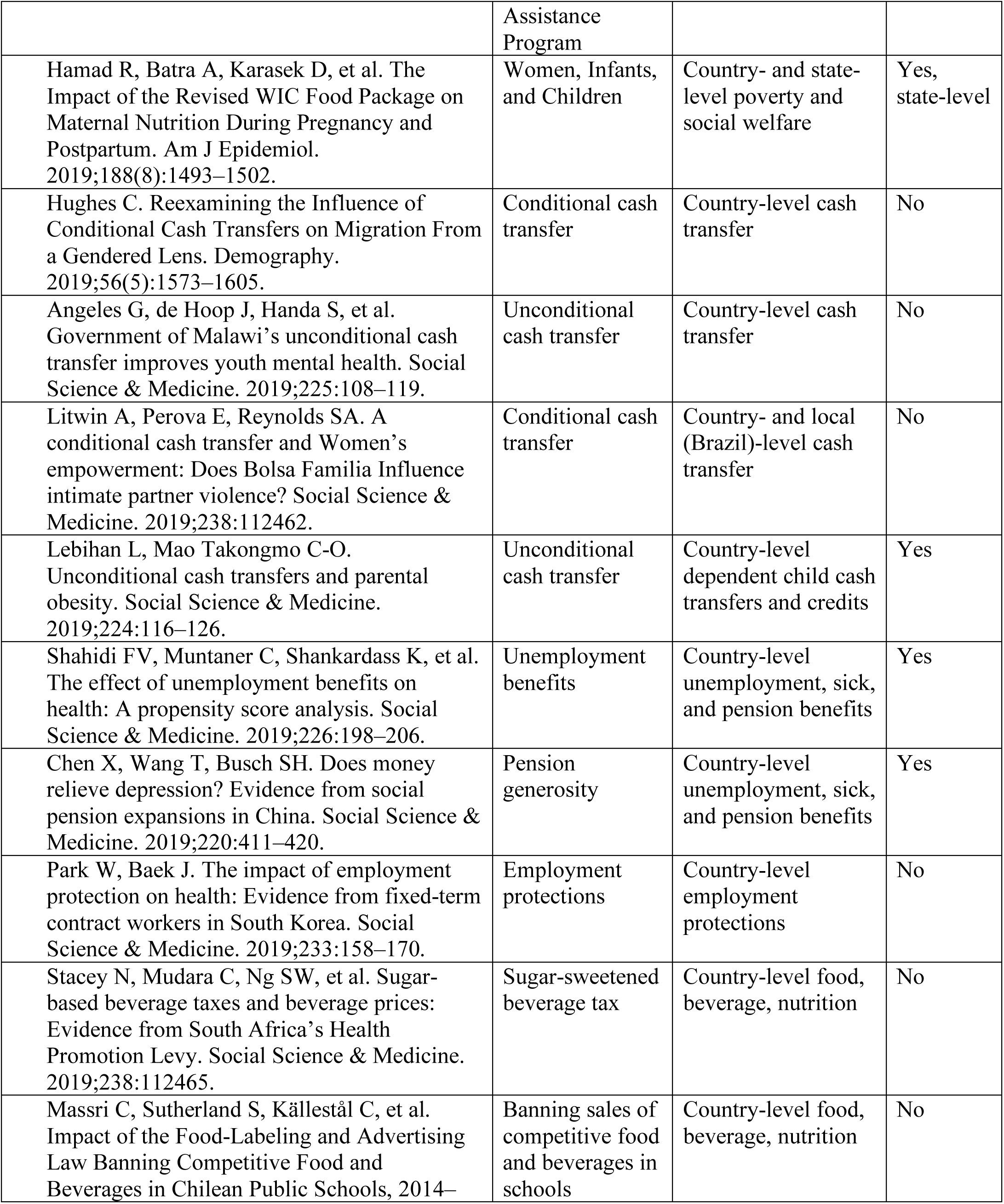
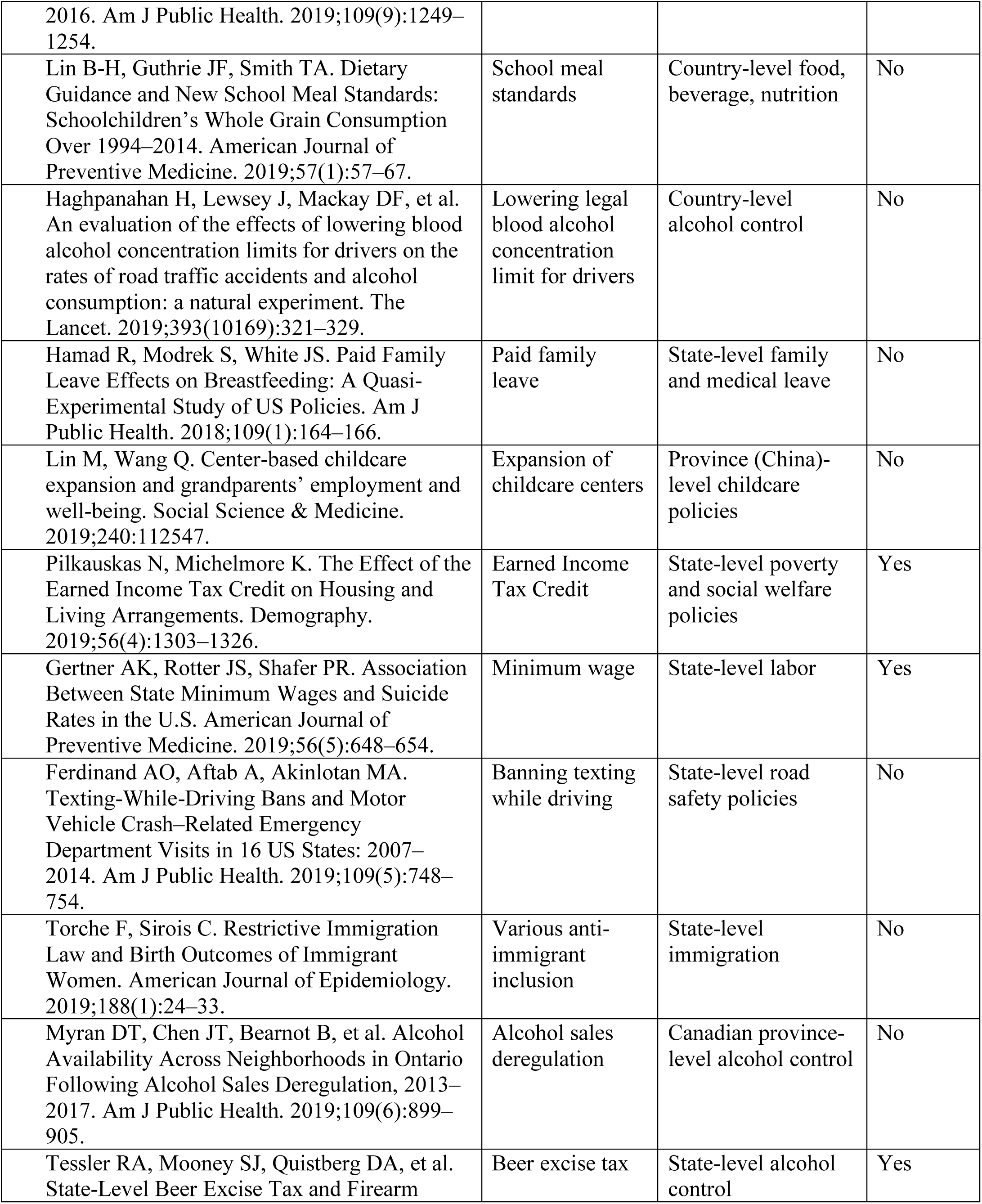
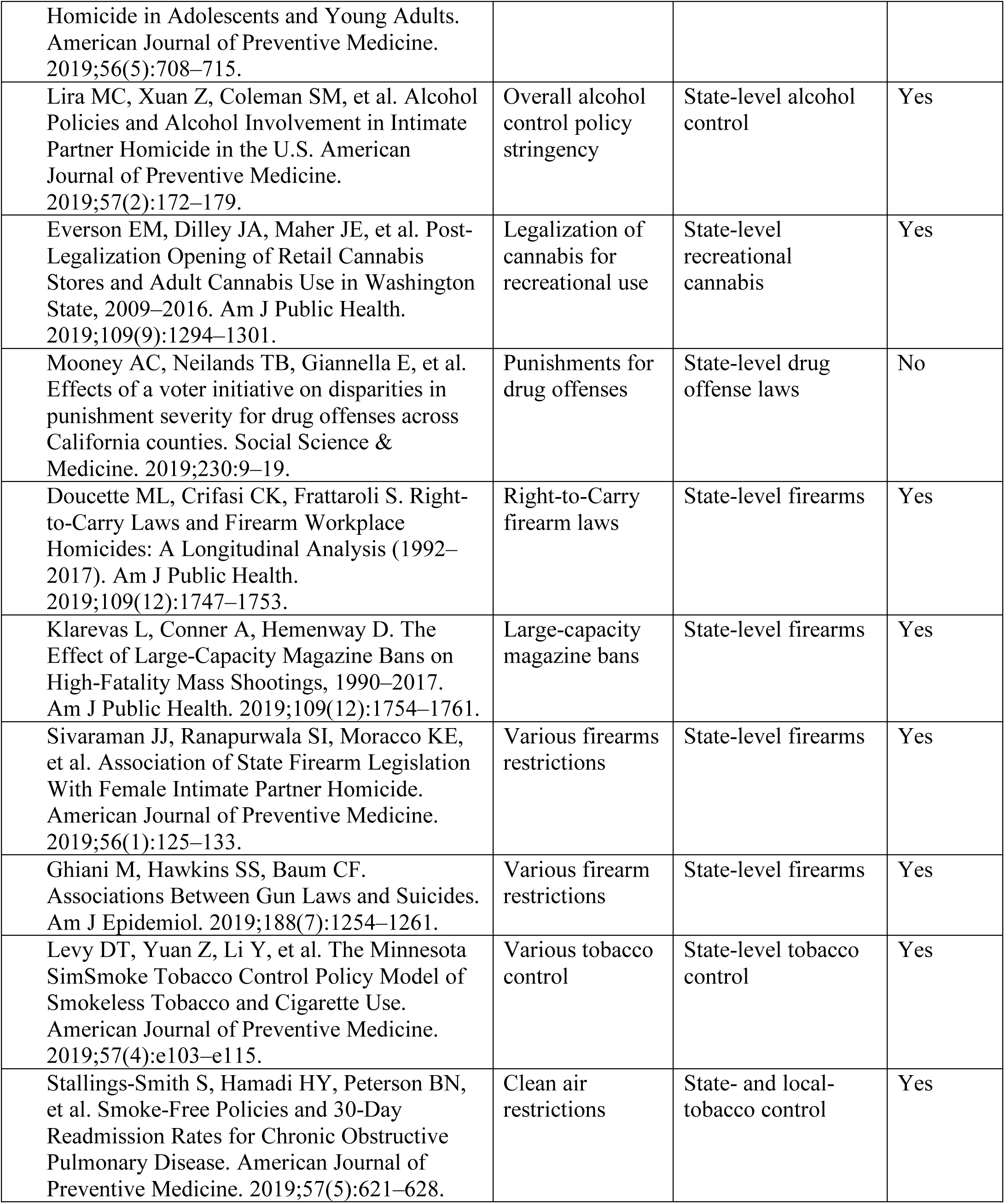
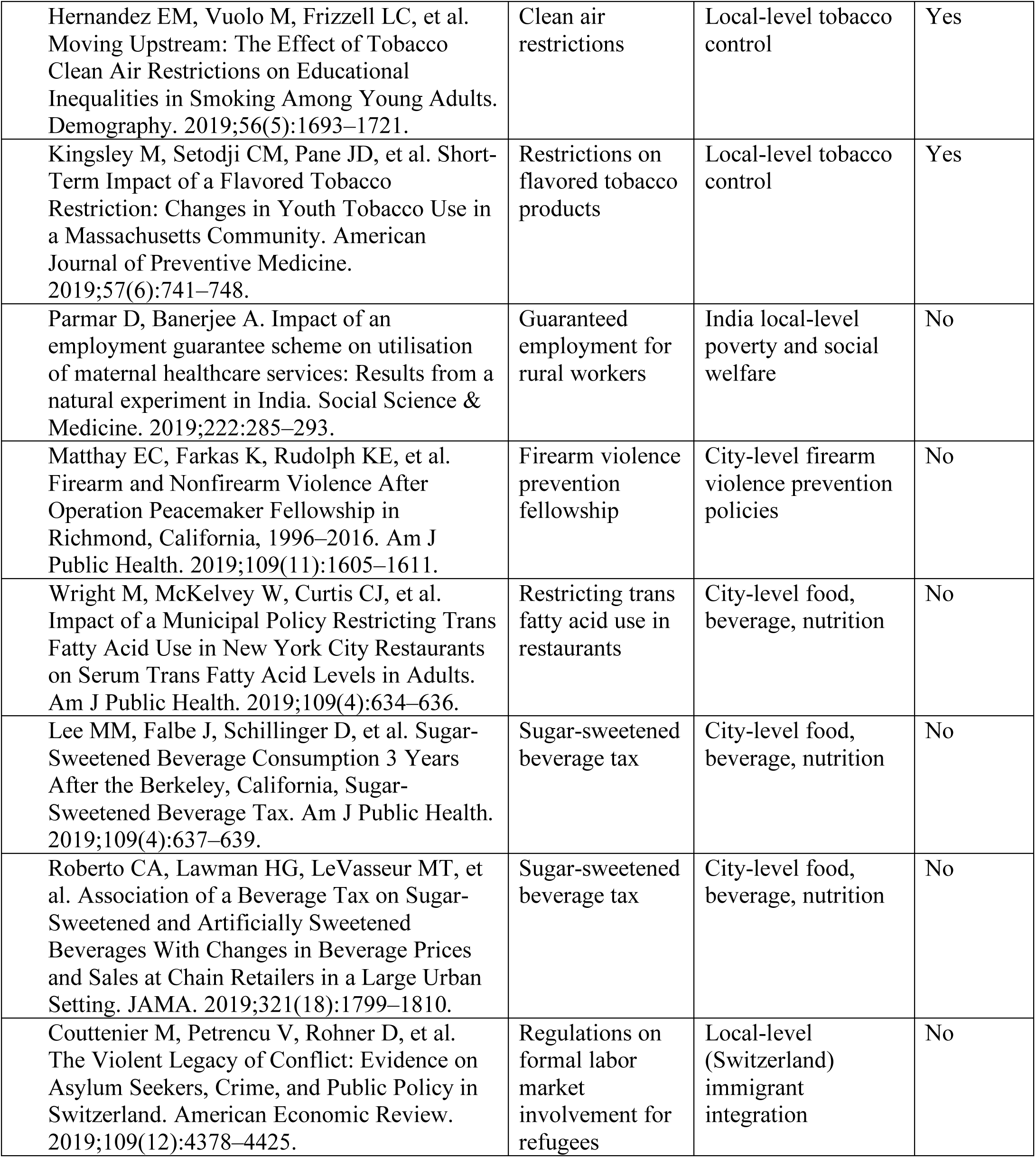
Social policy studies identified in systematic sample

**Appendix Table 2:** Variables dropped from statistical models due to strong correlations with other predictors

| **Model type** | **Database** | **Predictor policy variables per model (N, across models)** | **Predictor variables dropped (range, across models)** |
| --- | --- | --- | --- |
| Estimation of proportion of variance in each policy explained by other policies in the same database ($R^2$) | State poverty and social welfare | 10 – 10 | 0 - 0 |
|  | State labor and worker welfare | 25 - 25 | 0 - 0 |
|  | State firearms | 121 - 122 | 8 - 9 |
|  | State recreational cannabis | 22 - 23 | 7 - 8 |
|  | State alcohol control | 57 - 57 | 0 - 0 |
|  | State tobacco control | 32 - 33 | 1 - 2 |
|  | Country family leave | 97 - 98 | 2 - 3 |
|  | Country fertility and immigration | 29 - 29 | 0 - 0 |
|  | Country dependent child benefits | 5 - 5 | 0 - 0 |
|  | Country unemployment, sick, and pension benefits | 21 - 21 | 0 - 0 |
|  | Country LGBT rights | 40 - 40 | 0 - 0 |
|  | County tobacco control | 33 - 37 | 3 - 7 |
|  | City tobacco control | 39 - 40 | 0 - 1 |
| Estimation of impacts on precision (simulated outcome with panel fixed effects) | State poverty and social welfare | 11 – 11 | 0 - 0 |
|  | State labor and worker welfare | 26 - 26 | 0 - 0 |
|  | State firearms | 122 – 122 | 9 - 9 |
|  | State recreational cannabis | 23 - 23 | 8 - 8 |
|  | State alcohol control | 58 – 58 | 0 - 0 |
|  | State tobacco control | 33 - 33 | 2 - 2 |
|  | Country family leave | 98 – 98 | 3 - 3 |
|  | Country fertility and immigration | 30 - 30 | 0 - 0 |
|  | Country dependent child benefits | 6 – 6 | 0 - 0 |
|  | Country unemployment, sick, and pension benefits | 22 – 22 | 0 - 0 |
|  | Country LGBT rights | 41 - 41 | 0 - 0 |
|  | County tobacco control | 34 – 34 | 7 - 7 |
|  | City tobacco control | 33 - 33 | 8 - 8 |

**Appendix Figure 1:**
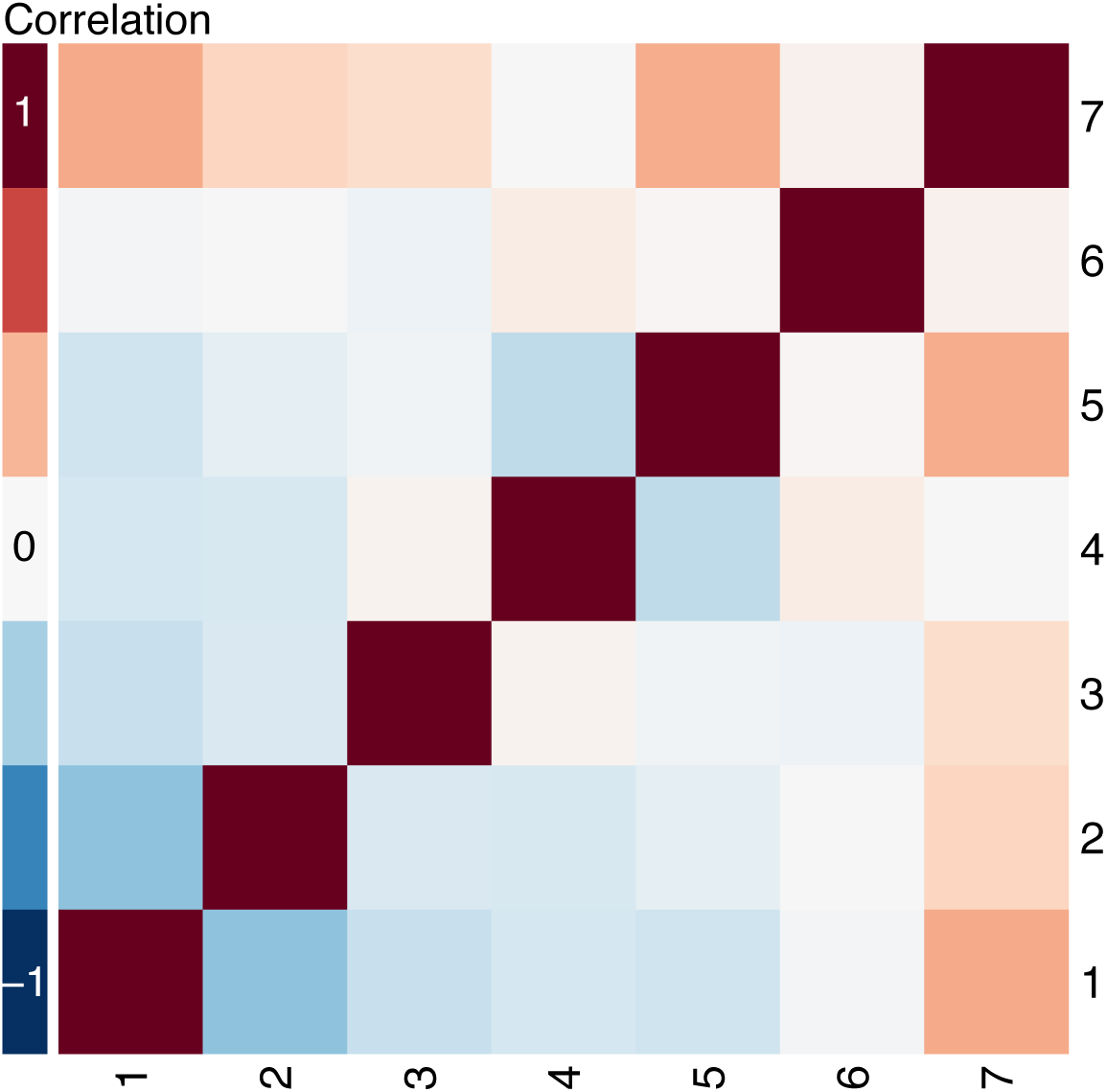
Heatmap of correlations among dependent child benefit policy measures across country-years, 35 countries, 1960-2015. Legend: High degrees of positive and negative correlation are indicated by the darkest red and blue colors, respectively. Policies: 1: Universal child benefit (amount). 2: Employment-based child benefit (amount). 3: Income-tested child benefit (amount). 4: Child tax allowance (amount). 5: Child tax credit (amount). 6: Child tax rebate (amount). 7: Total child benefit (amount).

**Appendix Figure 2:**
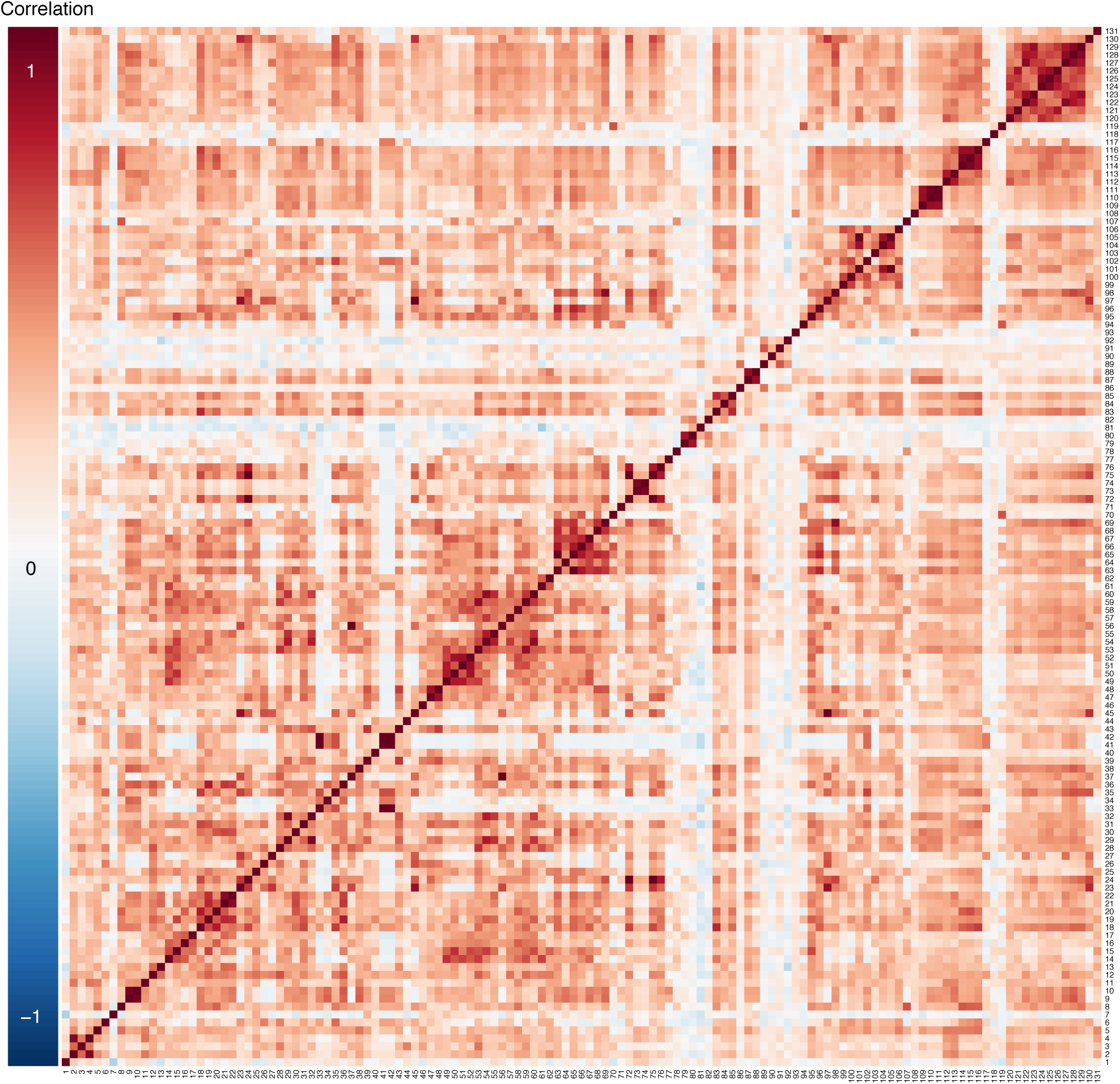
Heatmap of correlations among firearm policy measures across state-years, 50 states, 1991-2018. Legend: High degrees of positive and negative correlation are indicated by the darkest red and blue colors, respectively. Policies 1: Firearm possession is prohibited for all people with a felony conviction. 2: Firearm possession is prohibited for people who have been involuntarily committed to an inpatient facility. 3: Firearm possession is prohibited for people who have been involuntarily committed to an outpatient facility. 4: Firearm possession is prohibited if person is deemed by court to be a danger to oneself or others. 5: Firearm possession is prohibited for people with a drug misdemeanor conviction. 6: Firearm possession is prohibited for some people with alcohol-related problems. 7: Firearm possession is prohibited for some people with alcoholism. 8: People are required to relinquish their firearms after they become prohibited from possessing them. 9: Firearm possession is prohibited for people who have committed a violent misdemeanor punishable by less than one year of imprisonment. 10: Handgun possession is prohibited for people who have committed a violent misdemeanor punishable by less than one year of imprisonment. 11: Firearm possession is prohibited for people who have committed a violent misdemeanor punishable by more than one year of imprisonment. 12: State dealer license required for sale of all firearms. 13: State dealer license required for sale of handguns. 14: All private sellers and licensed dealers are required to keep and retain records of all firearm sales. 15: All private sellers and licensed dealers are required to keep and retain records of handgun sales. 16: Licensed dealers are required to keep and retain records of all firearm sales. 17: Licensed dealers are required to keep and retain records of handgun sales. 18: All private sellers and licensed dealers are required to report all firearm sales records to the state. 19: All private sellers and licensed dealers are required to report handgun sales records to the state. 20: Licensed dealers are required to report all firearm sales records to the state. 21: Licensed dealers are required to report handgun sales records to the state. 22: Dealers can retain sales records for at least 60 days after firearm purchase. 23: Ban on non-commercial dealers. 24: Mandatory reporting of stolen guns by all firearm dealers. 25: State requires at least one store security precaution for firearm dealers. 26: Mandatory police inspections of dealers. 27: Vendor license required to sell ammunition. 28: A license or permit is required to purchase all firearms. 29: A license or permit is required to purchase handguns. 30: Buyers must be fingerprinted at point of purchase. 31: Safety training or testing required prior to issuing a firearm license or permit. 32: Permit process involves law enforcement. 33: Gun owners must register their firearms with the state. 34: Gun owners must register their handguns with the state. 35: De facto registration of firearms is in place because of a recordkeeping requirement for all gun sales. 36: De facto registration of handguns is in place because of a recordkeeping requirement for all handgun sales. 37: Permit required to purchase ammunition. 38: All of the state’s high-risk gun possession prohibitions also apply to ammunition possession. 39: Purchase of handguns from licensed dealers and private sellers restricted to age 21 and older. 40: Purchase of long guns from licensed dealers and private sellers restricted to age 18 and older. 41: Purchase of long guns from licensed dealers restricted to age 21 and older. 42: Purchase of long guns from licensed dealers and private sellers restricted to age 21 and older. 43: No possession of handguns until age 21. 44: No possession of long guns until age 18. 45: No possession of long guns until age 21. 46: Mandatory reporting of lost and stolen guns by firearm owners. 47: Purchase of any type of ammunition restricted to age 18 and older. 48: Purchase of handgun ammunition restricted to age 21 and older. 49: Universal background checks required at point of purchase for all firearms. 50: Universal background check required at point of purchase for handguns. 51: Background checks required for all gun show firearm sales at point of purchase. 52: Background checks required for gun show handgun sales at point of purchase. 53: Background checks conducted through permit requirement for all firearm sales (or universal background checks). 54: Background checks conducted through permit requirement for all handgun sales (or universal background checks). 55: State can retain background check records for at least 60 days. 56: Background checks required for ammunition purchase. 57: Background checks for gun sales or permits have more than a three day period in which they can be completed. 58: Required background checks include an explicit requirement for search of state mental health records. 59: State conducts separate background checks, beyond the federal NICS check, for all firearms. 60: State conducts separate background checks, beyond the federal NICS check, for handguns. 61: Waiting period is required on all firearm purchases from dealers. 62: Waiting period is required on all handgun purchases from dealers. 63: Ban on sale of assault weapons beyond just assault pistols. 64: Ban on sale of assault weapons using a one-feature definition. 65: Ban on sale of assault weapons which includes a list of banned weapons. 66: Grandfathered weapons must be registered. 67: Transfer of grandfathered weapons is prohibited. 68: Ban on sale large capacity magazines beyond just ammunition for pistols. 69: No magazines with a capacity of more than 10 rounds of ammunition may be sold. 70: Possession of pre-owned large capacity magazines is prohibited. 71: Buyers can purchase no more than one handgun per month with no or limited exceptions. 72: No person may purchase a firearm with the intent to re-sell without the buyer going through a background check or having already gone through a background check. 73: No person may purchase a firearm with the intent to re-sell to a person who is prohibited from buying or possessing a firearm. 74: No person may purchase a handgun with the intent to re-sell to a person who is prohibited from buying or possessing a firearm. 75: No person may purchase a firearm on behalf of another person. 76: No person may purchase a handgun on behalf of another person. 77: All handguns sold must have either ballistic fingerprinting or microstamping so that they can be identified if used in a crime. 78: Law enforcement officers can confiscate firearms from any person who is deemed by a judge to represent a threat to themselves or others. 79: No gun carrying allowed on college campuses except for concealed weapon permittees. 80: No gun carrying on college campuses, including concealed weapons permittees. 81: No gun carrying on elementary school property, including concealed weapons permittees. 82: No open carry of handguns is allowed in public places. 83: No open carry of long guns is allowed in public places. 84: No open carry of handguns is allowed in public places unless the person has a concealed carry or handgun carry permit. 85: No open carry of long guns is allowed in public places unless the person has a permit. 86: Permit required to carry concealed weapons. 87: ℌMay issue” state. 88: Applicants are required to make a heightened showing to obtain a concealed carry permit. 89: Concealed carry permit process requires a background check. 90: Background check process for a concealed carry permit explicitly requires a check of the NICS database. 91: Concealed carry permit renewal requires a new background check. 92: Authorities are required to revoke concealed carry permits under certain circumstances. 93: No stand your ground law. 94: State has a law that requires review of personalized gun technology. 95: Safety lock required for handguns sold through licensed dealers. 96: Safety lock required for handguns sold through all dealers. 97: All firearms in a household must be stored securely (locked away) at all times. 98: Safety lock is required for handguns and must be approved by state standards. 99: Criminal liability for negligent storage of guns, regardless of whether child gains access. 100: Criminal liability for negligent storage of guns if child gains access. 101: Criminal liability for negligent storage of guns if child uses or carries the gun. 102: Criminal liability for negligent storage applies regardless of whether gun is loaded or unloaded. 103: Criminal liability for negligent storage applies to access by children less than 18 years old. 104: Criminal liability for negligent storage applies to access by children less than 16 years old. 105: Criminal liability for negligent storage applies to access by children less than 14 years old. 106: Ban on junk guns (sometimes called “Saturday night specials”). 107: Dealers are liable for damages resulting from illegal gun sales. 108: No law provides blanket immunity to gun manufacturers or prohibits state or local lawsuits against gun manufacturers. 109: State law does not preempt local regulation of firearms in any way. 110: Any state law that preempts local regulation of firearms is narrow in its scope (i.e., in one area of regulation). 111: State law does not completely preempt local regulation of firearms. 112: People convicted of a misdemeanor crime of domestic violence against a spouse, ex-spouse, or cohabitating partner are prohibited from possessing firearms. 113: All people convicted of a misdemeanor crime of domestic violence are prohibited from possessing firearms. 114: People convicted of a misdemeanor crime of domestic violence against a spouse, ex-spouse, or cohabitating partner are required to surrender their firearms. 115: People convicted of a misdemeanor crime of domestic violence against a spouse, ex-spouse, or cohabitating partner are required to surrender their firearms with no exceptions. 116: The surrender provisions apply if the defendant is a dating partner of the victim. 117: State law allows law enforcement to remove firearms from MCDV offenders. 118: State law requires law enforcement to remove firearms from the scene of a domestic violence incident. 119: All firearms must be removed from the scene of a domestic violence incident. 120: State law automatically prohibits domestic violence-related restraining order (DVRO) subjects from possessing firearms. 121: DVROs are automatically prohibiting if the subject is a dating partner of the petitioner. 122: Ex parte (temporary) DVRO subjects are automatically prohibited from possessing firearms. 123: Ex parte DVROs are prohibiting if the petitioner is a dating partner of the DVRO subject. 124: State law requires DVRO subjects to surrender their firearms. 125: No additional finding is required before the firearm surrender provisions apply. 126: The surrender provisions apply if the subject is a dating partner of the petitioner. 127: State law requires ex parte DVRO subjects to surrender their firearms. 128: No additional finding is required before the ex parte DVRO firearm surrender provisions apply. 129: The ex parte DVRO surrender provisions apply if the subject is a dating partner of the petitioner. 130: Law enforcement officials are required to remove firearms from people subject to a domestic violence-related restraining order. 131: A stalking conviction is prohibitive for firearm possession.

**Appendix Figure 3:**
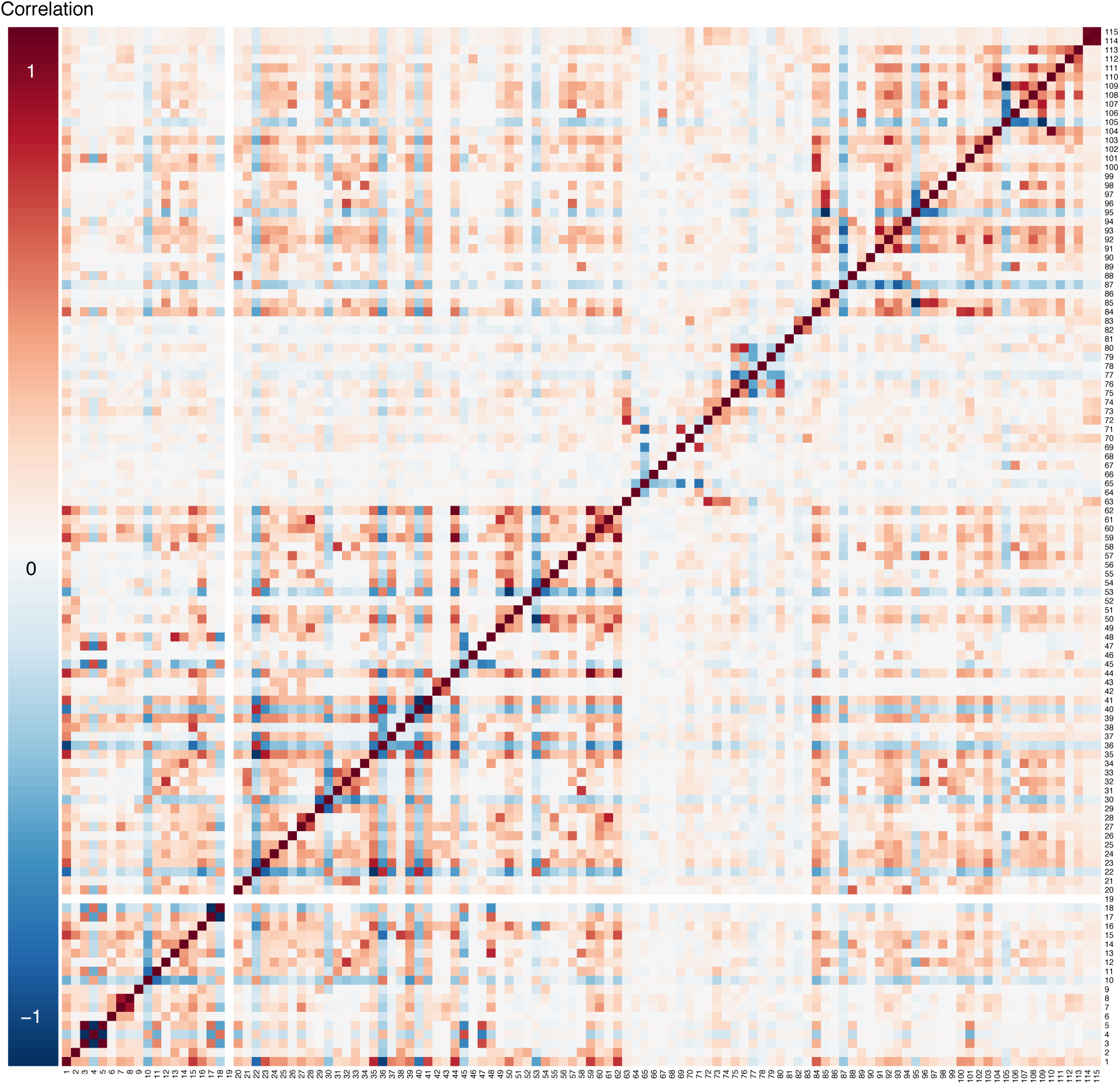
Heatmap of correlations among family leave-related policy measures across country-years, 190 countries, 1995-2016. Legend: High degrees of positive and negative correlation are indicated by the darkest red and blue colors, respectively. Policies: 1: Child health leave, any. 2: Child health leave for general health needs, maximum child age. 3: Child health leave for general health needs, rate depends on other reasons. 4: Child health leave for general health needs, rate has no variation. 5: Child health leave for general health needs, rate depends on number of children. 6: Child health leave for general health needs, duration. 7: Child health leave for general health needs requires minimum tenure and/or duration of contributions to social insurance fund, any. 8: Child health leave for general health needs requires minimum tenure and/or duration of contributions to social insurance fund, months required. 9: Child health leave for general health needs, depends on other reasons. 10: Child health leave for general health needs, length has no variation. 11: Child health leave for general health needs, length depends on child’s age. 12: Child health leave for general health needs, length depends on number of children. 13: Child health leave for general health needs, length depends on health needs of child with disability. 14: Child health leave for general health needs, length depends on marital status. 15: Child health leave for general health needs, wage replacement rate. 16: Child health leave for serious illness, maximum child age. 17: Child health leave for serious illness, rate depends on other reasons. 18: Child health leave for serious illness, rate has no variation. 19: Child health leave for serious illness, rate depends on severity of child illness. 20: Child health leave for serious illness, duration. 21: Child health leave for serious illness, other health conditions covered. 22: Child health leave for serious illness, health conditions covered. 23: Child health leave for serious illness, serious illness covered. 24: Child health leave for serious illness, disability covered. 25: Child health leave for serious illness, hospitalization covered. 26: Child health leave for serious illness, imminent death covered. 27: Child health leave for serious illness requires minimum tenure and/or duration of contributions to social insurance fund, any. 28: Child health leave for serious illness requires minimum tenure and/or duration of contributions to social insurance fund, months required. 29: Child health leave for serious illness, length depends on other reasons. 30: Child health leave for serious illness, length has no variation. 31: Child health leave for serious illness, length depends on child’s age. 32: Child health leave for serious illness, length depends on number of employee’s children. 33: Child health leave for serious illness, length depends on disability vs. illness. 34: Child health leave for serious illness, length depends on marital status. 35: Child health leave for serious illness, wage replacement rate. 36: No child health leave. 37: Child health leave for serious illness. 38: Child health leave for general health needs. 39: Child health leave for serious illness and general health needs. 40: Child health leave, available to women and men. 41: Child health leave, available to women only for serious health needs. 42: Child health leave, available to women only for general health needs. 43: Child health leave, available to women only for serious and general health needs. 44: Family health leave, any. 45: Family health leave, rate does not vary. 46: Family health leave, rate depends on severity of illness. 47: Family health leave, rate depens on illness vs. disability. 48: Family health leave, rate depends on other reasons. 49: Family health leave duration, duration. 50: Family health leave, limited to special cases such as serious illness or hospitalization. 51: Family health leave for general illness and special cases. 52: Family health leave, limited to other cases. 53: Family health leave, not limited to particular cases. 54: Family health leave, limited to serious illness. 55: Family health leave, limited to disability. 56: Family health leave, limited to hospitalization. 57: Family health leave, limited to imminent death. 58: Family health leave, limited to when spouse is sick. 59: Family health leave, only for specific family members and/or with a same-residency requirement, any. 60: Family health leave requires minimum tenure and/or duration of contributions to social insurance fund, any. 61: Family health leave requires minimum tenure and/or duration of contributions to social insurance fund, months required. 62: Family health leave, wage replacement rate. 63: Maternity leave, any. 64: Maternity leave, rate depends on other reasons. 65: Maternity leave, rate has no variation. 66: Maternity leave, rate depends on worker tenure. 67: Maternity leave, rate depends on income level. 68: Maternity leave, rate depends on child’s order among other children. 69: Maternity leave, rate depends on duration of leave. 70: Maternity leave requires minimum contribution to social insurance fund, any. 71: Maternity leave, maximum wage replacement rate. 72: Maternity leave, minimum wage replacement rate. 73: Maternity leave, paid weeks. 74: Maternity leave, maximum weeks before birth. 75: Maternity leave, weeks required after birth. 76: Maternity leave, weeks required before birth. 77: Maternity leave, no required set durations for pre-birth, post-birth, or both. 78: Maternity leave, required set durations for pre-birth. 79: Maternity leave, required set durations for post-birth. 80: Maternity leave, required set durations for pre- and post-birth. 81: Maternity leave, required set durations for pre-birth, post-birth, or both, but unspecified. 82: Maternity leave requires minimum tenure and/or duration of contributions to social insurance fund, any. 83: Maternity leave requires minimum tenure and/or duration of contributions to social insurance fund, months required. 84: Parental leave, any. 85: Parental leave, any incentives for men to take. 86: Parental leave, rate depends on other reasons. 87: Parental leave, rate has no variation. 88: Parental leave, rate depends on worker tenure. 89: Parental leave, rate depends on income level. 90: Parental leave, rate depends on child’s order among other children. 91: Parental leave, rate depends on duration of leave. 92: Parental leave requires minimum contribution to social insurance fund, any. 93: Parental leave, maximum wage replacement rate. 94: Parental leave, maximum weeks paid. 95: Parental leave, no incentive for men. 96: Parental leave, incentive for men that leave cannot be transferred to partner. 97: Parental leave, incentive of bonus if both parents take some leave. 98: Parental leave, incentive that benefit is reduced if father does not take minimum leave. 99: Parental leave required for men. 100: Parental leave, minimum wage replacement rate. 101: Parental leave, minimum weeks paid. 102: Parental leave requires minimum tenure and/or duration of contributions to social insurance fund, any. 103: Parental leave requires minimum tenure and/or duration of contributions to social insurance fund, months required. 104: Paternity leave, any. 105: Paternity leave, rate has no variation. 106: Paternity leave, rate depends on income level. 107: Paternity leave, rate depends on duration of leave. 108: Paternity leave requires minimum contribution to social insurance fund, any. 109: Paternity leave, maximum wage replacement rate. 110: Paternity leave, minimum wage replacement rate. 111: Paternity leave, paid weeks. 112: Paternity leave requires minimum tenure and/or duration of contributions to social insurance fund, any. 113: Paternity leave requires minimum tenure and/or duration of contributions to social insurance fund, months required. 114: Sick leave, any. 115: Sick leave is paid.

**Appendix Figure 4:**
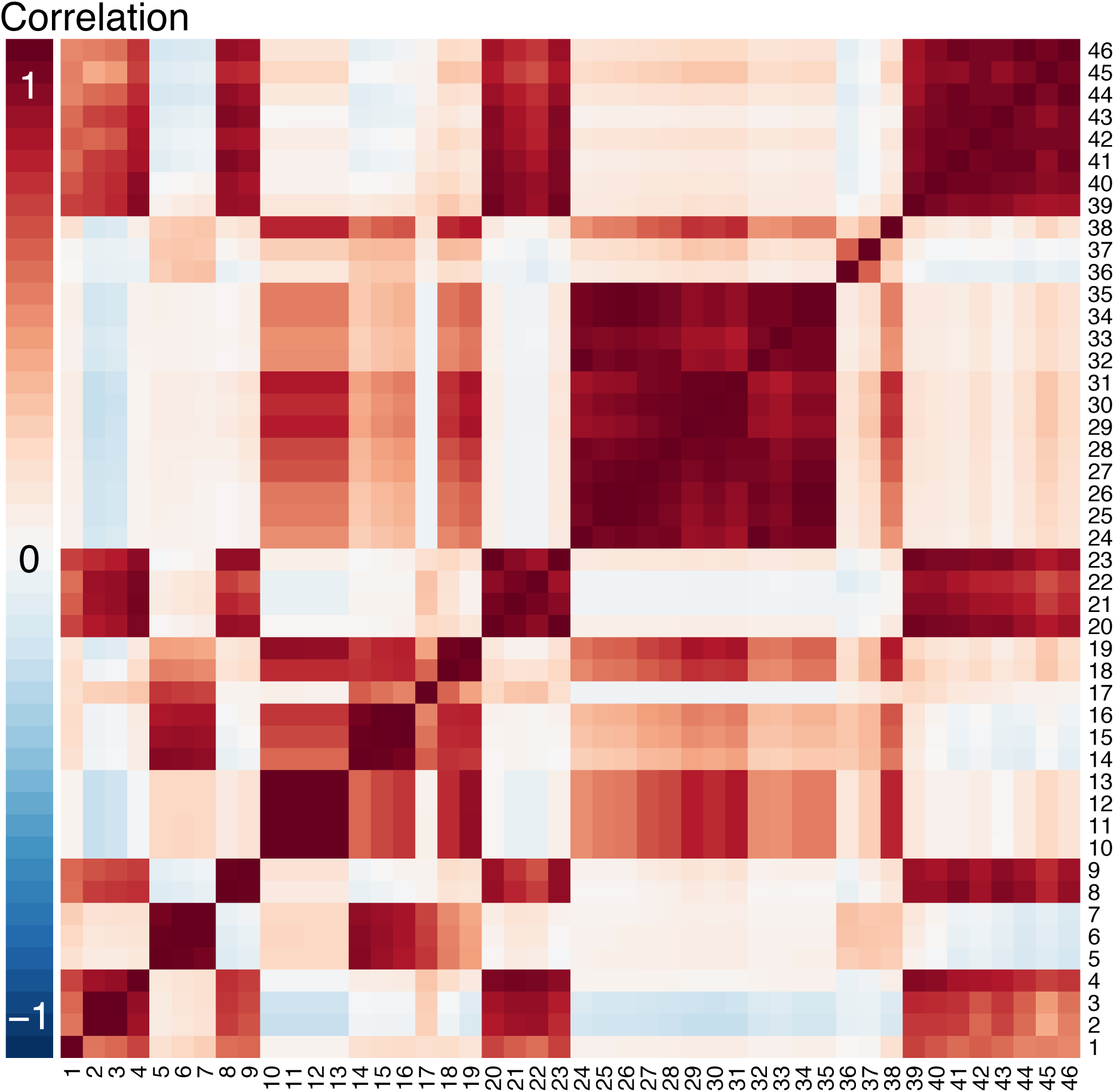
Heatmap of correlations among poverty and social welfare policy measures across state-years, 50 states, 1990-2011. Legend: High degrees of positive and negative correlation are indicated by the darkest red and blue colors, respectively. Abbreviations: AFDC: Aid to Families with Dependent Children. TANF: Temporary Assistance to Needy Families. SNAP: Supplemental Nutrition Assistance Program. FS: Food Stamps. SSI: Supplemental Security Income. EITC: Earned Income Tax Credit. WIC: Special Supplemental Nutrition Program for Women, Infants, and Children. NSLP: National School Lunch Program. SBP: School Breakfast Program. Policies: 1: Workers compensation. 2: AFDC/TANF recipients. 3: AFDC/TANF caseloads. 4: Child-only AFDC/TANF cases. 5: AFDC/TANF benefit for 2-person family. 6: AFDC/TANF benefit for 3-person family. 7: AFDC/TANF benefit for 4-person family. 8: FS/SNAP Recipients. 9: FS/SNAP caseloads. 10: FS/SNAP benefit for 1-person family. 11: FS/SNAP benefit for 2-person family. 12: FS/SNAP benefit for 3-person family. 13: FS/SNAP benefit for 4-person family. 14: AFDC/TANF/FS 2-person benefit. 15: AFDC/TANF/FS 3-person benefit. 16: AFDC/TANF/FS 4-person benefit. 17: State SSI. 18: Total SSI. 19: SSI/FS Benefit. 20: SSI recipients. 21: SSI recipients who are aged. 22: SSI recipients who are blind. 23: SSI recipients who are disabled. 24: EITC phase-in rate, no dependents. 25: EITC phase-in rate, 1 dependent. 26: EITC phase-in rate, 2 dependents. 27: EITC phase-in rate, 3 dependents. 28: EITC maximum credit, no dependents. 29: EITC maximum credit, 1 dependent. 30: EITC maximum credit, 2 dependents. 31: EITC maximum credit, 3 dependents. 32: EITC phase-out rate, no dependents. 33: EITC phase-out rate, 1 dependent. 34: EITC phase-out rate, 2 dependents. 35: EITC phase-out rate, 3 dependents. 36: State EITC rate. 37: Refundable state EITC. 38: State minimum wage. 39: Medicaid beneficiaries. 40: WIC participants. 41: NSLP, free participants. 42: NSLP, reduced participants. 43: NSLP, total participants. 44: SBP, free participants. 45: SBP, reduced participants. 46: SBP, total participants.

**Appendix Figure 5:**
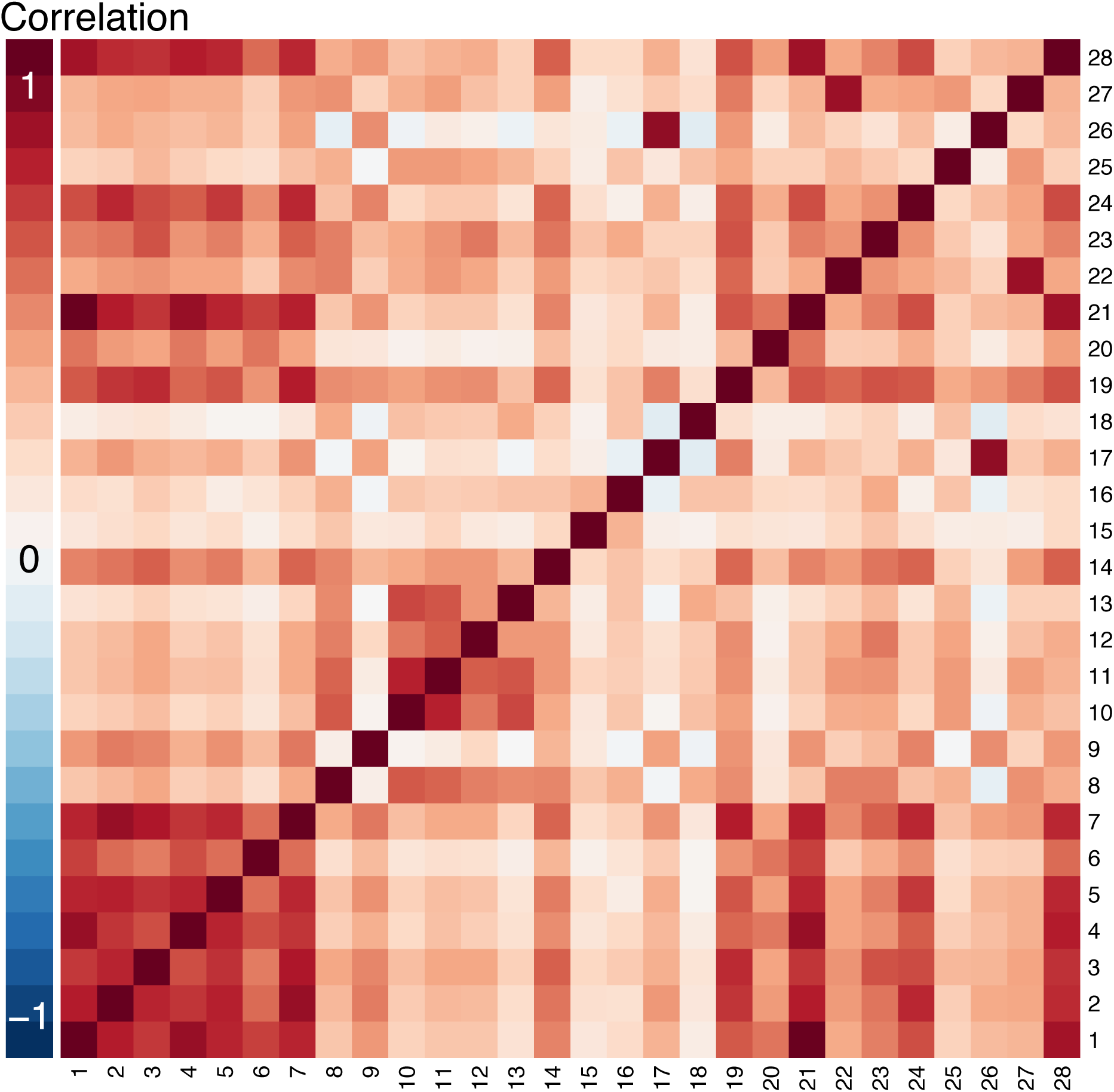
Heatmap of correlations among labor policy measures across state-years, 50 states, 1865-2015. Legend: High degrees of positive and negative correlation are indicated by the darkest red and blue colors, respectively. Policies: 1: Accountant licensing, source 1. 2: Architects licensing. 3: Beautician licensing. 4: Child labor standards. 5: Chiropractors licensing. 6: Dentists licensing. 7: Engineer licensing. 8: Fair employment laws. 9: Integrated bar. 10: Collective bargaining rights for state employees. 11: Collective bargaining rights for local teachers. 12: Minimum wage law for men. 13: State minimum wage law above federal level. 14: Prevailing wage law. 15: Labor relations law patterned after Taft-Hartley. 16: Labor relations law patterned after Wagner Act. 17: Right to work law, source 1. 18: State temporary disability insurance program. 19: State pension system. 20: State labor agency. 21: Accountant licensing, source 2. 22: Migratory labor committee. 23: Minimum wage law. 24: Real estate broker licensing. 25: Retainers agreement. 26: Right to work law, source 2. 27: Seasonal agricultural labor standards. 28: Workmen’s compensation.

**Appendix Figure 6:**
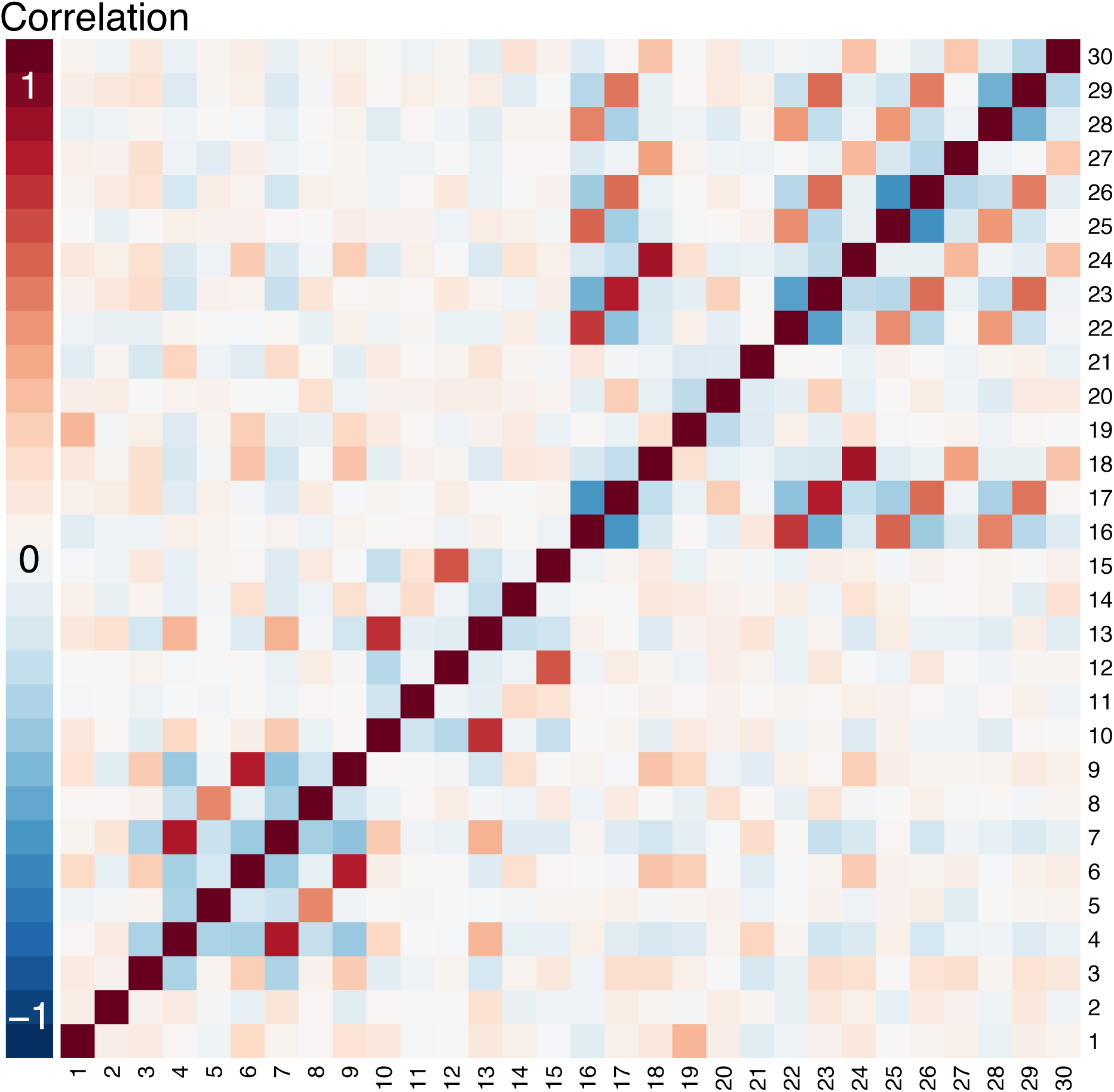
Heatmap of correlations among fertility and immigration policy measures across country-years, 199 countries, 1996-2011. Legend: High degrees of positive and negative correlation are indicated by the darkest red and blue colors, respectively. Policies: 1: Encourage the return of citizens. 2: Has policy on adolescent fertility. 3: Has policy on integrating non-nationals. 4: Policy to lower population growth. 5: Policy to maintain population growth. 6: Policy to raise population growth. 7: Policy to lower fertility. 8: Policy to maintain fertility. 9: Policy to raise fertility. 10: Policy to lower rural-urban migration. 11: Policy to maintain rural-urban migration. 12: Policy to raise rural-urban migration. 13: Policy to lower migration into urban agglomerations. 14: Policy to maintain migration into urban agglomerations. 15: Policy to raise migration into urban agglomerations. 16: Policy to lower immigration. 17: Policy to maintain immigration. 18: Policy to raise immigration. 19: Policy to lower emigration. 20: Policy to maintain emigration. 21: Policy to increase emigration. 22: Policy to lower permanent settlement. 23: Policy to maintain permanent settlement. 24: Policy to raise permanent settlement. 25: Policy to lower temporary workers. 26: Policy to maintain temporary workers. 27: Policy to raise temporary workers. 28: Policy to lower family reunification. 29: Policy to maintain family reunification. 30: Policy to raise family reunification.

**Appendix Figure 7:**
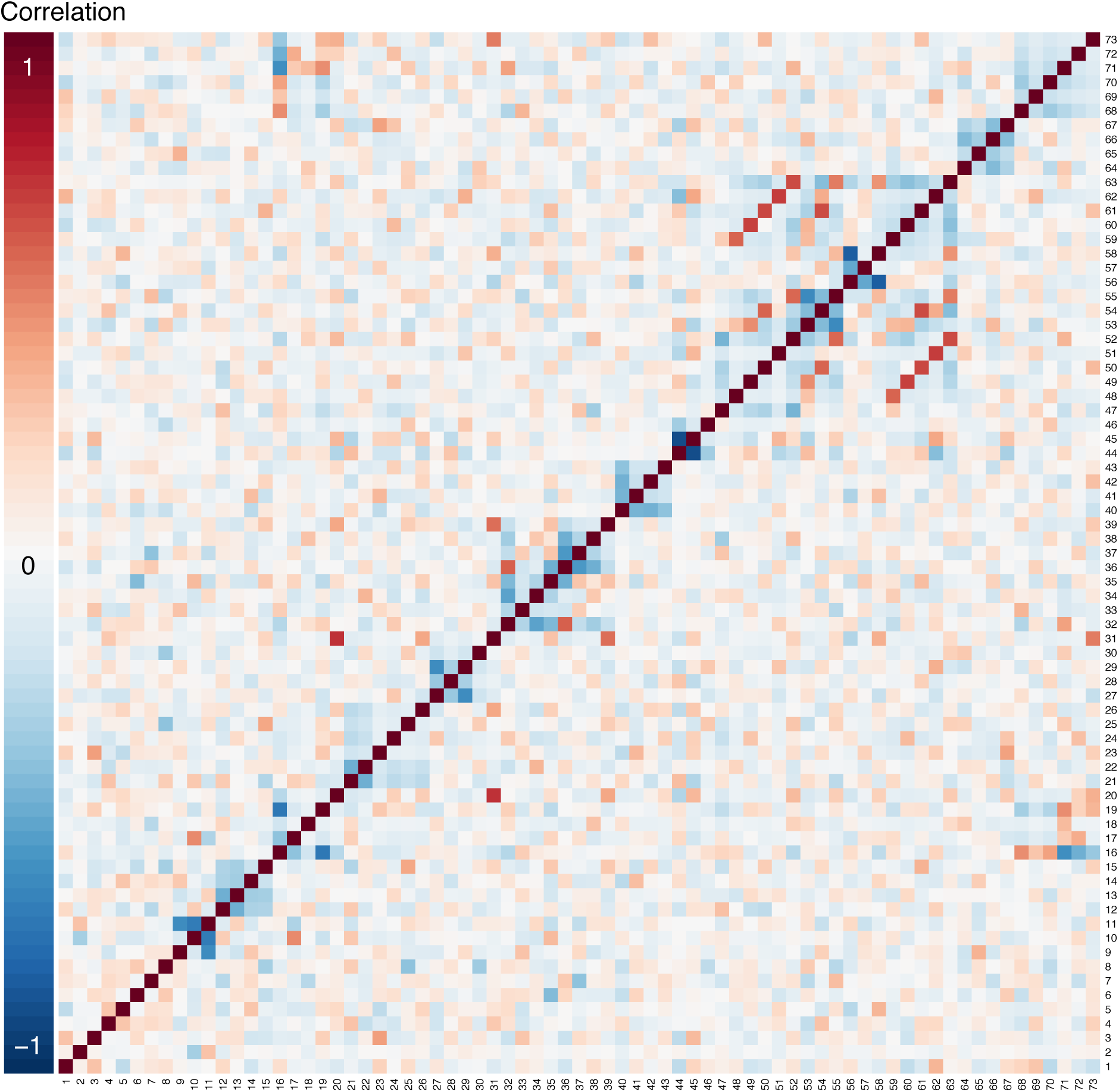
Heatmap of correlations among alcohol control policy measures across state-years, 50 states, 2003-2018. Legend: High degrees of positive and negative correlation are indicated by the darkest red and blue colors, respectively. Policies: 1: Off-premise beer taxes. 2: Adult blood alcohol concentration limit. 3: Fake ID, retailer has no affirmative defense. 4: Fake ID, retailer can seize. 5: Fake ID, supplier liable. 6: Fake ID, user liable. 7: Open containers in motor vehicles prohibited. 8: Underage purchase prohibited. 9: Boating blood alcohol concentration limit .08 or .10, no per se. 10: Boating blood alcohol concentration limit .10, per se. 11: Boating blood alcohol concentration limit .08, per se. 12: No responsible beverage service training. 13: Responsible beverage service training voluntary. 14: Responsible beverage service training mandatory for some. 15: Responsible beverage service training mandatory for all. 16: All private distribution of wine, beer, and spirits. 17: Partial or complete state control of wholesale for wine and spirits. 18: Private and state wine and spirits stores; most wholesale wine private; wholesale spirits are state controlled. 19: All or most spirits are state controlled; most wine private for stores and wholesale. 20: All wine and spirits sold only in state stores and state wholesale. 21: No restrictions on drink specials. 22: Restricts some drink specials but not happy hour. 23: Restricts some happy hours. 24: Restricts happy hours and unlimited beverages for fixed price. 25: Bans happy hour. 26: Bans happy hour and unlimited beverages for fixed price. 27: No keg registration. 28: Keg registration but lacking key components. 29: Keg registration with 2 key components. 30: Keg registration with 3 key components. 31: Kegs banned. 32: Sellers for off-premise can be under 21 and without supervision. 33: Sellers for off-premise beer can be under 21 and without supervision; sellers of off-premise spirits have to be 21. 34: Sellers for off-premise can be under 21 but with supervision. 35: Sellers for off-premise must be 21. 36: Sellers for on-premise can be under 21 and without supervision. 37: Sellers for on-premise can be under 21 and without supervision but not bartenders. 38: Sellers for on-premise can be under 21 but with supervision. 39: Sellers for on-premise and bartenders must be 21. 40: No social host law. 41: Social host, sub-threshold. 42: Social host, meets threshold. 43: Social host, threshold plus. 44: Sunday sales allowed. 45: Some prohibitions on Sunday sales. 46: Sunday sales prohibited. 47: No law on underage consumption. 48: Lax law on underage consumption. 49: Intermediate 1 law on underage consumption. 50: Intermediate 2 law on underage consumption. 51: Intermediate 3 law on underage consumption. 52: Underage consumption prohibited, no exceptions. 53: Parent/guardian can furnish. 54: Parent/guardian can furnish in private locations. 55: Furnishing prohibited, no exceptions. 56: No underage internal possession law. 57: Underage internal possession allowed with requirements. 58: Underage internal possession banned, no exceptions. 59: Underage possession allowed in private locations. 60: Underage possession allowed with parental consent. 61: Underage possession allowed in private locations with parental consent. 62: Underage possession allowed in parent’s home. 63: Underage possession prohibited, no exceptions. 64: No use-lose law. 65: Discretionary authority use-lose. 66: Mandatory authority use-lose. 67: Use-lose with 4 key components. 68: No policies on wholesale price. 69: 1 of 3 restrictions on wholesale price of 1-2 beverage types. 70: 1 of 3 restrictions on wholesale price of all beverage types. 71: All 3 restrictions on wholesale price of spirits. 72: All 3 restrictions on wholesale price of spirits and wine; 1 of 3 restrictions on wholesale price of beer. 73: All 3 restrictions on wholesale price of spirits and wine; 2-3 of 3 restrictions on wholesale price of beer.

**Appendix Figure 8:**
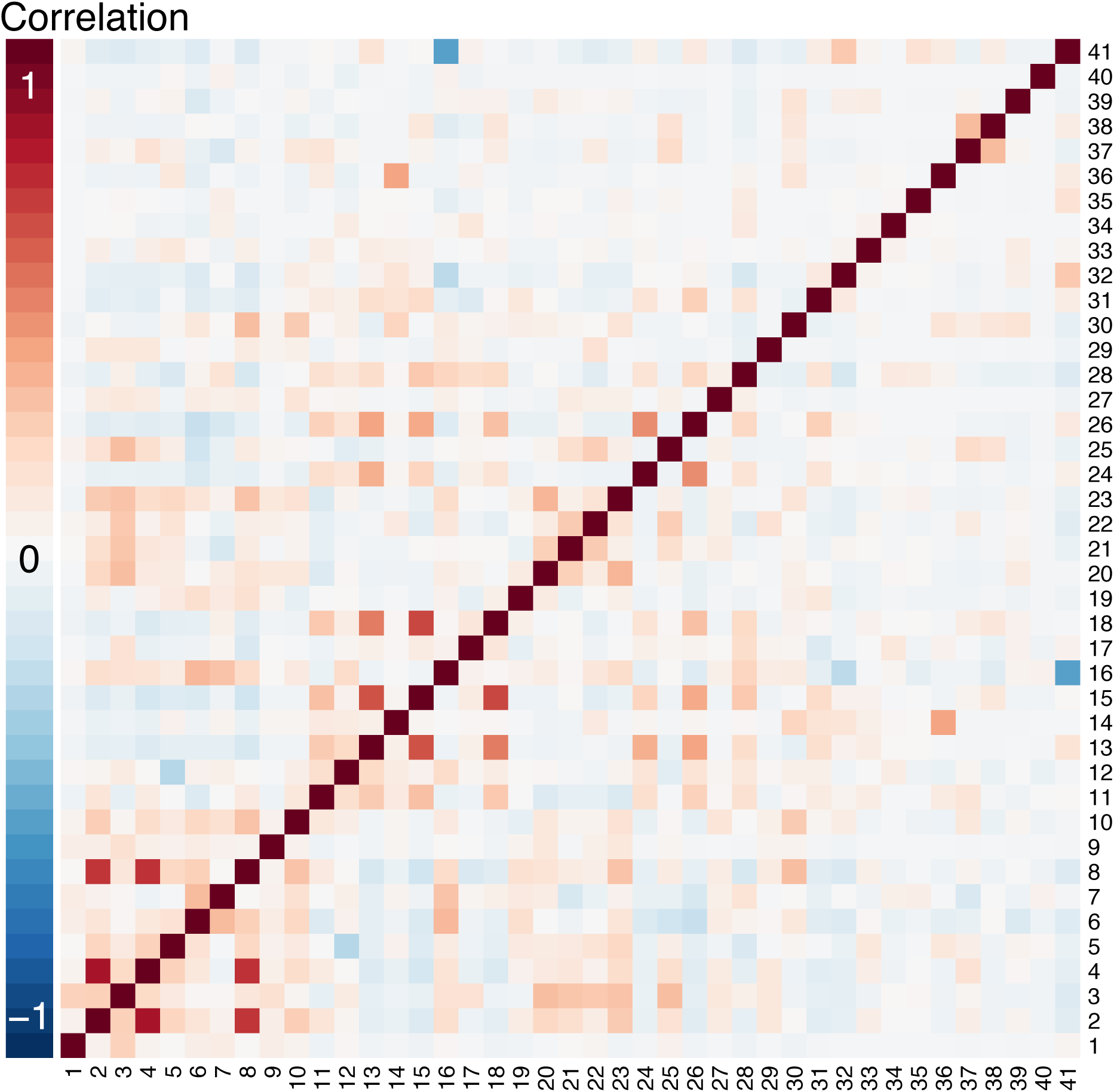
Heatmap of correlations among LGBT rights policy measures across country-months, 229 countries, 1729-2020. Legend: High degrees of positive and negative correlation are indicated by the darkest red and blue colors, respectively. Abbreviations: LGBT: lesbian, gay, bisexual, and transgender. Policies: 1: Homosexual activity, illegal death penalty as punishment. 2: LBGT discrimination, no protections. 3: Same-sex marriage, not legal. 4: LBGT employment discrimination, no protections. 5: Unequal age of consent. 6: Same-sex adoption, single only. 7: Homosexuals serving openly in military, legal. 8: LGBT housing discrimination, no protections. 9: Homosexual activity, male illegal, female uncertain. 10: Same-sex marriage, unrecognized. 11: Homosexual activity, legal. 12: Equal age of consent. 13: LGBT discrimination illegal. 14: LGBT employment discrimination, sexual orientation protected only. 15: LGBT housing discrimination protection for sexual orientation and gender identity only. 16: Blood donations by men who have sex with men, legal. 17: Conversion therapy not banned. 18: LGBT employment discrimination, protection for sexual orientation and gender identity. 19: Same-sex marriage, foreign same-sex marriages recognized only. 20: Homosexual activity, illegal imprisonment as punishment. 21: Homosexuals serving openly in military, illegal. 22: Right to change legal gender, illegal. 23: Homosexual activity, male illegal, female legal. 24: Same-sex marriage, legal. 25: Same-sex adoption, illegal. 26: Same-sex adoption, legal. 27: Homosexual activity, illegal, other penalty. 28: Right to change legal gender, legal surgery not required. 29: Homosexual activity, illegal, up to life in prison as punishment. 30: LGBT discrimination illegal in some contexts. 31: Conversion therapy banned. 32: Right to change legal gender, legal but requires surgery. 33: Same-sex marriage, civil unions. 34: Same-sex adoption, stepchild adoption only. 35: Same-sex marriage, unregistered cohabitation. 36: LGBT housing discrimination protection for sexual orientation only. 37: Homosexuals serving openly in military, don’t ask, don’t tell. 38: Same-sex marriage unrecognized, same-sex marriage and civil unions banned. 39: Same-sex adoption, married couples only. 40: Same-sex marriage unrecognized, only same-sex marriage official. 41: Blood donations by men who have sex with men banned.

**Appendix Figure 9:**
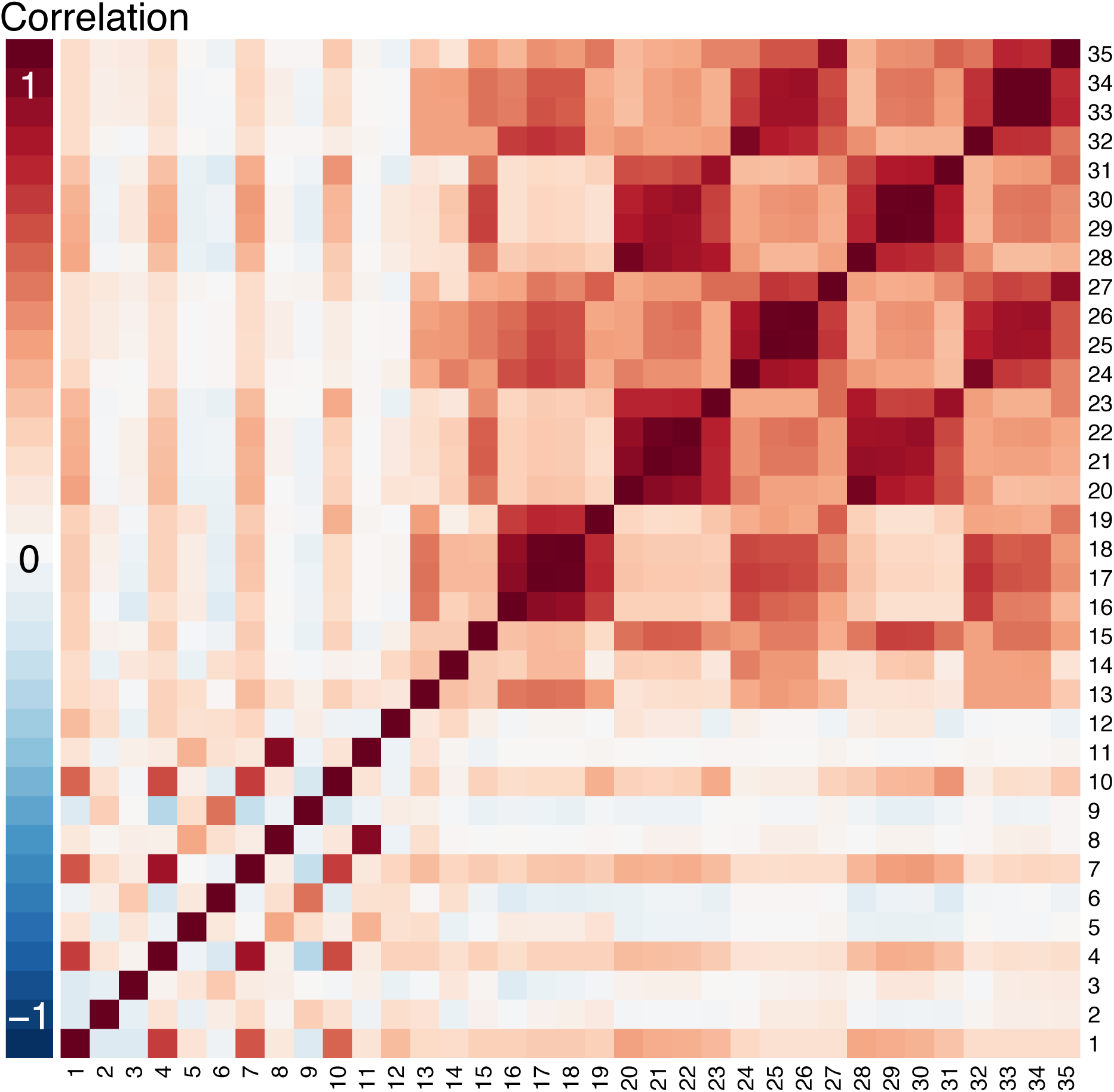
Heatmap of correlations between policy measures across state-months in database of state tobacco control policies, 55 states and territories, June 2003 – December 2020. Legend: High degrees of positive and negative correlation are indicated by the darkest red and blue colors, respectively. Policies: 1: 100% clean air in workplaces. 2: Qualified clean air law for workplaces. 3: Some clean air provisions for workplaces. 4: 100% clean air in restaurants. 5: Qualified clean air law for restaurants. 6: Some clean air provisions for restaurants. 7: 100% clean air in bars. 8: Qualified clean air law for bars. 9: Some clean air provisions for bars. 10: 100% clean air in gaming facilities. 11: Qualified clean air law for gaming facilities. 12: Some clean air provisions for gaming facilities. 13: Clean air law addresses e-cigarettes. 14: Clean air law addresses hookah. 15: Clean air law addresses marijuana. 16: 100% clean air in workplaces, includes e-cigarettes. 17: 100% clean air in restaurants, includes e-cigarettes. 18: 100% clean air in bars, includes e-cigarettes. 19: 100% clean air in gaming facilities, includes e-cigarettes. 20: 100% clean air in workplaces, includes recreational marijuana smoke. 21: 100% clean air in restaurants, includes recreational marijuana smoke. 22: 100% clean air in bars, includes recreational marijuana smoke. 23: 100% clean air in gaming facilities, includes recreational marijuana smoke. 24: 100% clean air in workplaces, includes recreational marijuana vaping. 25: 100% clean air in restaurants, includes recreational marijuana vaping. 26: 100% clean air in bars, includes recreational marijuana vaping. 27: 100% clean air in gaming facilities, includes recreational marijuana vaping. 28: 100% clean air in workplaces, includes medical marijuana smoke. 29: 100% clean air in restaurants, includes medical marijuana smoke. 30: 100% clean air in bars, includes medical marijuana smoke. 31: 100% clean air in gaming facilities, includes medical marijuana smoke. 32: 100% clean air in workplaces, includes medical marijuana vaping. 33: 100% clean air in restaurants, includes medical marijuana vaping. 34: 100% clean air in bars, includes medical marijuana vaping. 35: 100% clean air in gaming facilities, includes medical marijuana vaping.

**Appendix Figure 10:**
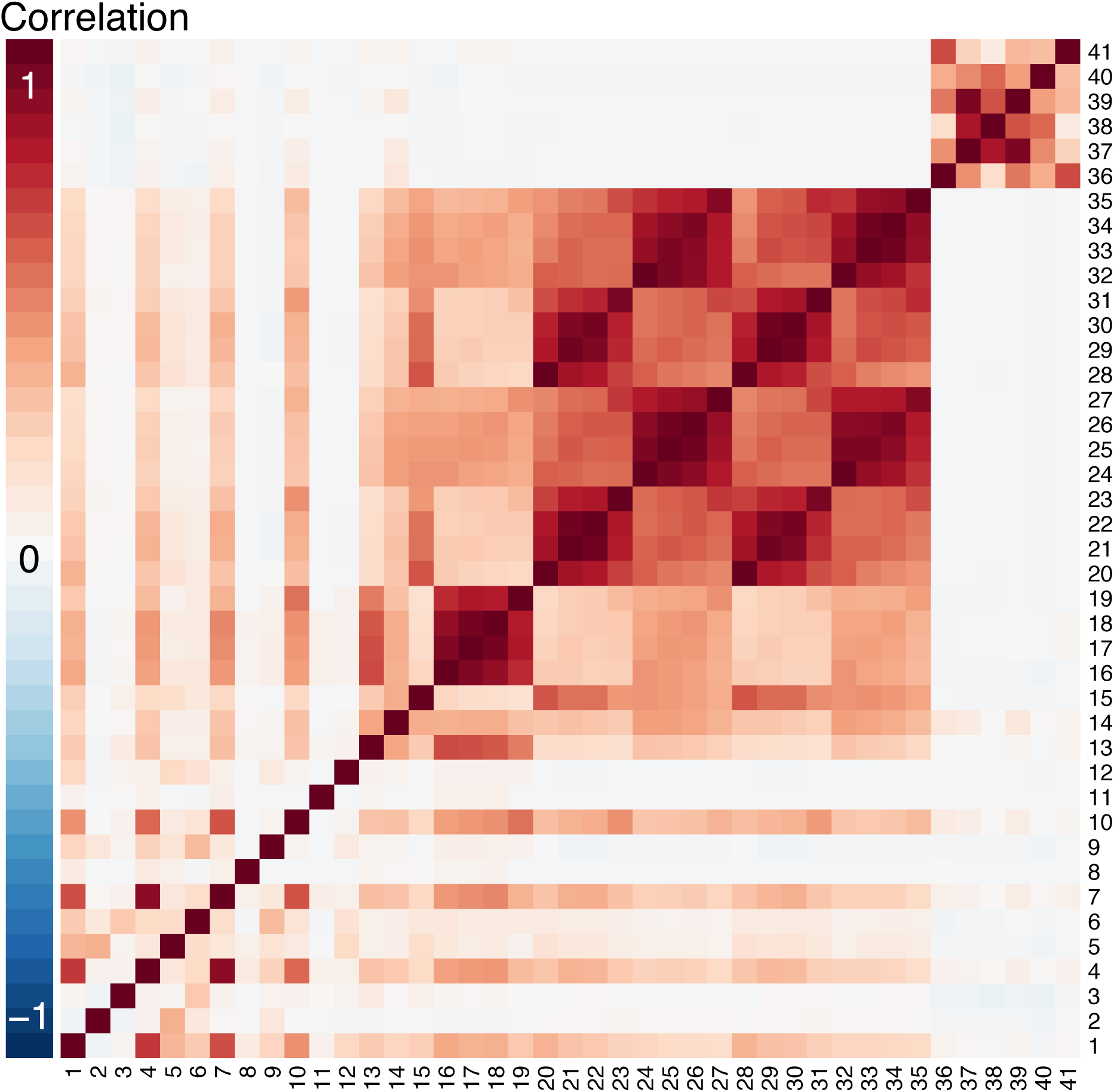
Heatmap of correlations between policy measures across county-months in database of county tobacco control policies, 772 counties, October 1986 – December 2020. Legend: High degrees of positive and negative correlation are indicated by the darkest red and blue colors, respectively. Policies: 1: 100% clean air in workplaces. 2: Qualified clean air law for workplaces. 3: Some clean air provisions for workplaces. 4: 100% clean air in restaurants. 5: Qualified clean air law for restaurants. 6: Some clean air provisions for restaurants. 7: 100% clean air in bars. 8: Qualified clean air law for bars. 9: Some clean air provisions for bars. 10: 100% clean air in gaming facilities. 11: Qualified clean air law for gaming facilities. 12: Some clean air provisions for gaming facilities. 13: Clean air law addresses e-cigarettes. 14: Clean air law addresses hookah. 15: Clean air law addresses marijuana. 16: 100% clean air in workplaces, includes e-cigarettes. 17: 100% clean air in restaurants, includes e-cigarettes. 18: 100% clean air in bars, includes e-cigarettes. 19: 100% clean air in gaming facilities, includes e-cigarettes. 20: 100% clean air in workplaces, includes recreational marijuana smoke. 21: 100% clean air in restaurants, includes recreational marijuana smoke. 22: 100% clean air in bars, includes recreational marijuana smoke. 23: 100% clean air in gaming facilities, includes recreational marijuana smoke. 24: 100% clean air in workplaces, includes recreational marijuana vaping. 25: 100% clean air in restaurants, includes recreational marijuana vaping. 26: 100% clean air in bars, includes recreational marijuana vaping. 27: 100% clean air in gaming facilities, includes recreational marijuana vaping. 28: 100% clean air in workplaces, includes medical marijuana smoke. 29: 100% clean air in restaurants, includes medical marijuana smoke. 30: 100% clean air in bars, includes medical marijuana smoke. 31: 100% clean air in gaming facilities, includes medical marijuana smoke. 32: 100% clean air in workplaces, includes medical marijuana vaping. 33: 100% clean air in restaurants, includes medical marijuana vaping. 34: 100% clean air in bars, includes medical marijuana vaping. 35: 100% clean air in gaming facilities, includes medical marijuana vaping. 36: Excise taxes on cigarettes. 37: Excise taxes on cigars. 38: Excise taxes on little cigars. 39: Excise taxes on non-combustible tobacco. 40: Excise taxes on other combustible tobacco products. 41: Excise taxes on nicotine delivery devices or e-cigarette products.

**Appendix Figure 11:**
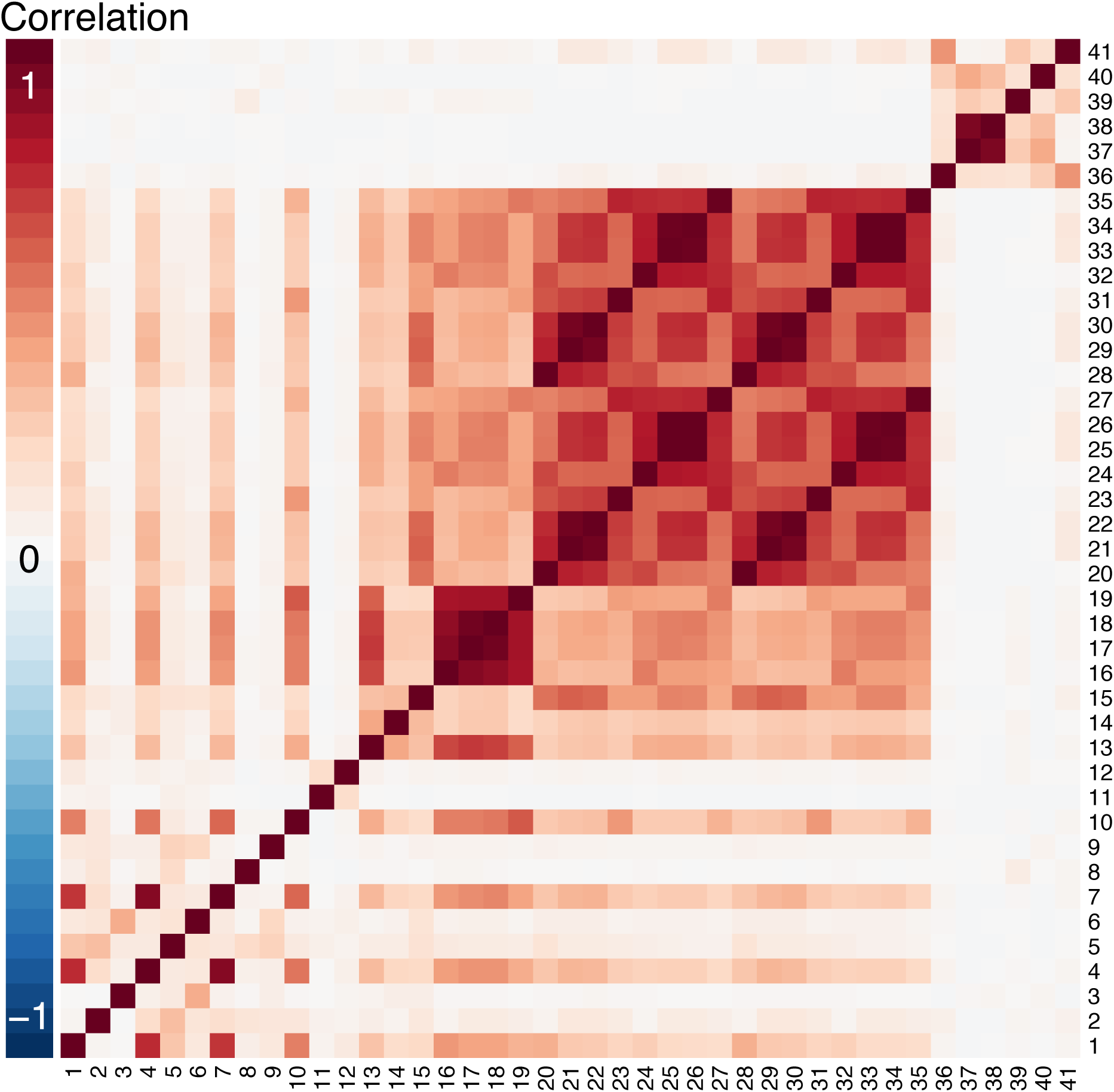
Heatmap of correlations between policy measures across city-months in database of city tobacco control policies, 3,204 cities, October 1986 – December 2020. Legend: High degrees of positive and negative correlation are indicated by the darkest red and blue colors, respectively. Policies: 1: 100% clean air in workplaces. 2: Qualified clean air law for workplaces. 3: Some clean air provisions for workplaces. 4: 100% clean air in restaurants. 5: Qualified clean air law for restaurants. 6: Some clean air provisions for restaurants. 7: 100% clean air in bars. 8: Qualified clean air law for bars. 9: Some clean air provisions for bars. 10: 100% clean air in gaming facilities. 11: Qualified clean air law for gaming facilities. 12: Some clean air provisions for gaming facilities. 13: Clean air law addresses e-cigarettes. 14: Clean air law addresses hookah. 15: Clean air law addresses marijuana. 16: 100% clean air in workplaces, includes e-cigarettes. 17: 100% clean air in restaurants, includes e-cigarettes. 18: 100% clean air in bars, includes e-cigarettes. 19: 100% clean air in gaming facilities, includes e-cigarettes. 20: 100% clean air in workplaces, includes recreational marijuana smoke. 21: 100% clean air in restaurants, includes recreational marijuana smoke. 22: 100% clean air in bars, includes recreational marijuana smoke. 23: 100% clean air in gaming facilities, includes recreational marijuana smoke. 24: 100% clean air in workplaces, includes recreational marijuana vaping. 25: 100% clean air in restaurants, includes recreational marijuana vaping. 26: 100% clean air in bars, includes recreational marijuana vaping. 27: 100% clean air in gaming facilities, includes recreational marijuana vaping. 28: 100% clean air in workplaces, includes medical marijuana smoke. 29: 100% clean air in restaurants, includes medical marijuana smoke. 30: 100% clean air in bars, includes medical marijuana smoke. 31: 100% clean air in gaming facilities, includes medical marijuana smoke. 32: 100% clean air in workplaces, includes medical marijuana vaping. 33: 100% clean air in restaurants, includes medical marijuana vaping. 34: 100% clean air in bars, includes medical marijuana vaping. 35: 100% clean air in gaming facilities, includes medical marijuana vaping. 36: Excise taxes on cigarettes. 37: Excise taxes on cigars. 38: Excise taxes on little cigars. 39: Excise taxes on non-combustible tobacco. 40: Excise taxes on other combustible tobacco products. 41: Excise taxes on nicotine delivery devices or e-cigarette products.

### R statistical code for simulating variance inflation due to policy co-occurrence

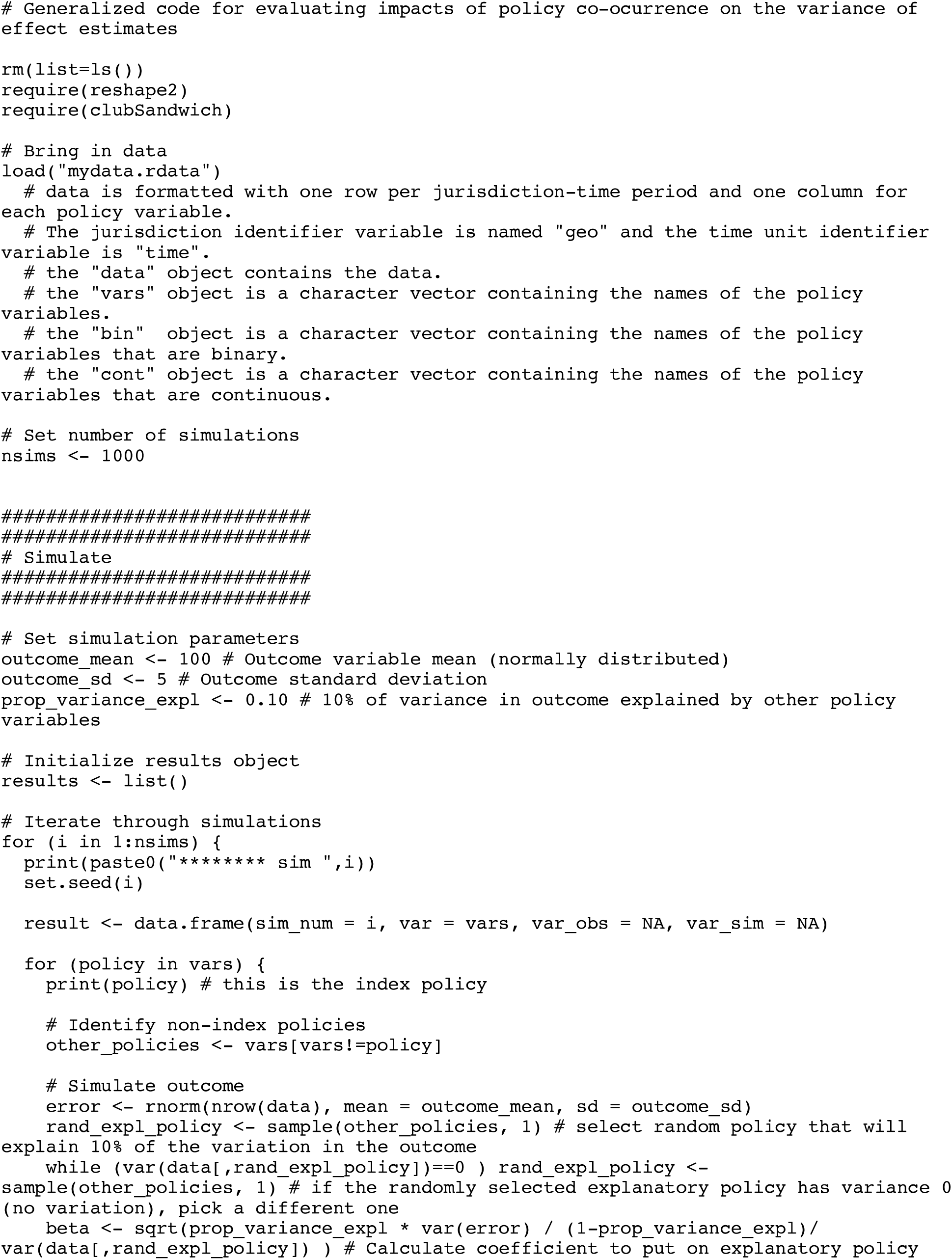
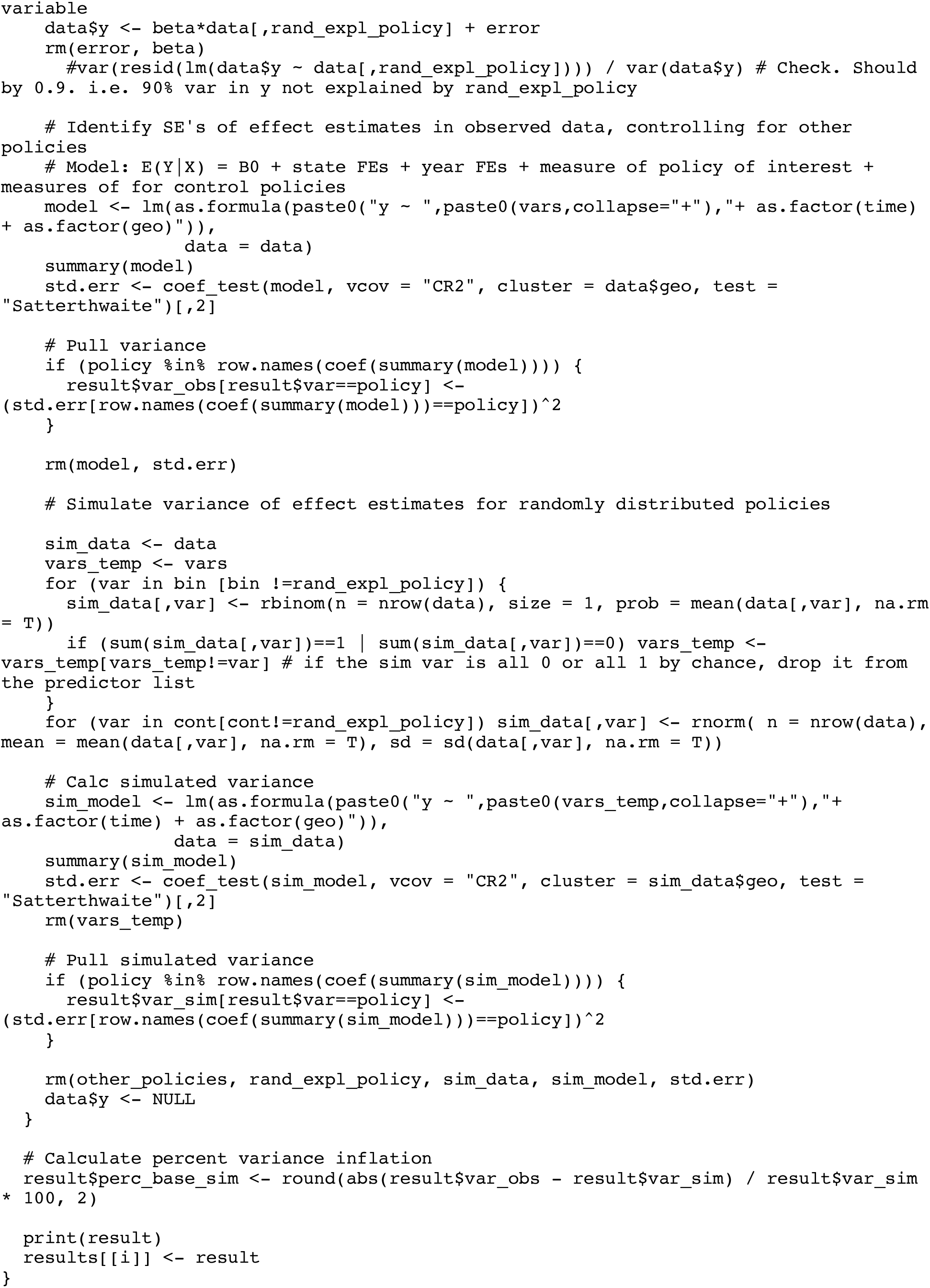
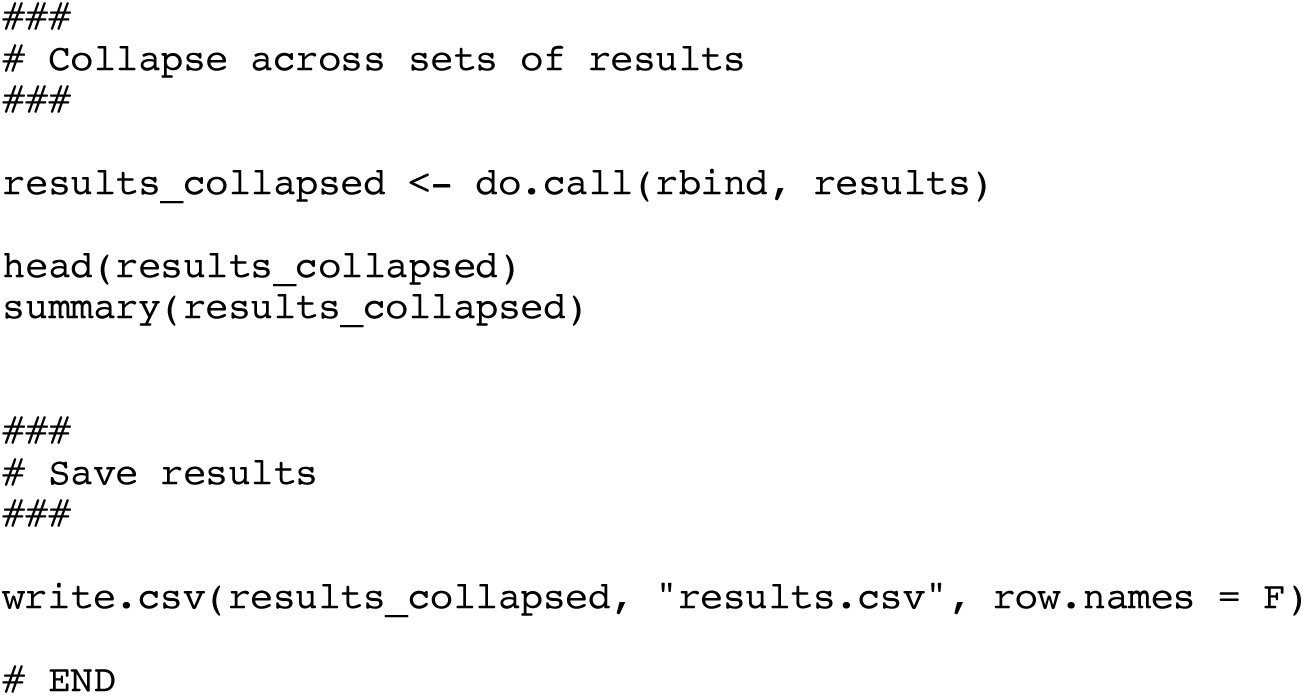

